# Phenotype Scoring of Population Scale Single-Cell Data Dissects Alzheimer’s Disease Complexity

**DOI:** 10.1101/2024.11.01.24316586

**Authors:** Chenfeng He, Athan Z. Li, Kalpana Hanthanan Arachchilage, Chirag Gupta, Xiang Huang, Xinyu Zhao, PsychAD Consortium, Kiran Girdhar, Georgios Voloudakis, Gabriel E. Hoffman, Jaroslav Bendl, John F. Fullard, Donghoon Lee, Panos Roussos, Daifeng Wang

**Affiliations:** Department of Biostatistics and Medical Informatics, University of Wisconsin-Madison, Madison, WI, 53706, USA; Waisman Center, University of Wisconsin-Madison, Madison, WI, 53705, USA; Department of Computer Sciences, University of Wisconsin-Madison, Madison, WI, 53706, USA; Department of Neuroscience, University of Wisconsin-Madison, Madison, WI, USA; PsychAD Consortium; Center for Disease Neurogenomics, Icahn School of Medicine at Mount Sinai, New York, NY, 10029, USA; Friedman Brain Institute, Icahn School of Medicine at Mount Sinai, New York, NY, 10029, USA; Department of Psychiatry, Icahn School of Medicine at Mount Sinai, New York, NY, 10029, USA; Department of Genetics and Genomic Science, Icahn School of Medicine at Mount Sinai, New York, NY, 10029, USA; Center for Precision Medicine and Translational Therapeutics, James J. Peters VA Medical Center, Bronx, NY, 10468, USA; Mental Illness Research Education and Clinical Center, James J. Peters VA Medical Center, Bronx, NY, 10468, USA

## Abstract

The complexity of Alzheimer’s disease (AD) manifests in diverse clinical phenotypes, including cognitive impairment and neuropsychiatric symptoms (NPSs). However, the etiology of these phenotypes remains elusive. To address this, the PsychAD project generated a population-level single-nucleus RNA-seq dataset comprising over 6 million nuclei from the prefrontal cortex of 1,494 individual brains, covering a variety of AD-related phenotypes that capture cognitive impairment, severity of pathological lesions, and the presence of NPSs. Leveraging this dataset, we developed a deep learning framework, called Phenotype Associated Single Cell encoder (PASCode), to score single-cell phenotype associations, and identified ∼1.5 million phenotype associate cells (PACs). We compared PACs within 27 distinct brain cell subclasses and prioritized cell subpopulations and their expressed genes across various AD phenotypes, including the upregulation of a reactive astrocyte subtype with neuroprotective function in AD resilient donors. Additionally, we identified PACs that link multiple phenotypes, including a subpopulation of protoplasmic astrocytes that alter their gene expression and regulation in AD donors with depression. Uncovering the cellular and molecular mechanisms underlying diverse AD phenotypes has the potential to provide valuable insights towards the identification of novel diagnostic markers and therapeutic targets. All identified PACs, along with cell type and gene expression information, are summarized into an AD-phenotypic single-cell atlas for the research community.

## Main

Alzheimer’s disease (AD) is a complex, progressive neurodegenerative disorder, which is characterized by a combination of pathological, cognitive, and neuropsychiatric symptoms (NPSs) that affect millions of individuals worldwide^1^. Due to the heterogeneity of AD, the disease can be characterized by various phenotypes, including cognitive decline, NPSs, and neuropathological lesions, which vary based on the stage of disease progression. Furthermore, some individuals with AD pathologies are cognitively normal and are referred to as ‘resilient’ to AD^2^. In addition, the role of NPSs in AD is understudied^3^ but they are important clinical features of AD progression associated with accelerated cognitive impairment^4^. These different phenotypes are categorized as high-level physiological abnormalities, the causes of which can usually be traced back to molecular alterations of cells^5^. Thus, pinpointing the cell states (subpopulation), genes and gene modules that are associated with certain AD phenotypes could facilitate the uncovering of disease mechanisms, prognostic gene biomarkers and novel therapeutic targets^2,6^.

The PsychAD Consortium generated population-level single-nucleus RNA sequencing data (snRNA-seq) consisting of more than 6.3 million nuclei isolated from the dorsolateral prefrontal cortex (DLPFC) of 1,494 donors^7^. The DLPFC is a key brain region for AD, for instance, it can be impaired in AD patients with memory loss^8^. The PsychAD cohort represents a variety of AD-related phenotypes, including Braak stages that measure progression neurofibrillary tangle pathology^9^, Clinical Dementia Rating (CDR)^10^, resilience to AD and a range of NPSs (such as depression and agitation) that capture the heterogeneity of AD across different stages of the disease. By inferring cell subpopulations and gene expression patterns associated with the aforementioned phenotypes, this dataset provides a unique opportunity to dissect the cellular and molecular mechanisms underlying AD.

To accomplish this, we developed the Phenotype Associated Single Cell encoder (PASCode), which is a deep learning framework that identifies phenotype associated cells (PACs). For robust inference, this framework ensembles several computational methods to detect cell subpopulations with differential abundance across phenotypes **(Supplementary Table 1)**^11,12^. Although those methods have been used to improve identification of disease related genes and pathways in, for example, COVID-19 and cancer^11,13^, they have not been widely applied to study AD. Therefore, after benchmarking and showing the outperformance of PASCode compared to existing methods, we applied it to the PsychAD dataset to infer PACs associated with various AD phenotypes in order to dissect the cellular and molecular etiology of AD. We present our results in four major sections. First, AD associated PACs were identified, followed by prioritization of specific glial (e.g., microglia, astrocyte) and neuronal subpopulations important for AD. Second, we identified PACs associated with the severity of AD progression, as well as cell subpopulations, such as reactive astrocytes, intratelencephalic excitatory neurons and SST inhibitory neurons that appear to confer resilience to AD. Third, we prioritized several cell subpopulations, such as protoplasmic astrocytes, that reveal the potential contribution of inflammation and Endoplasmic Reticulum stress pathways to depression in AD donors. Furthermore, we performed gene expression, cell trajectory and gene regulatory network analyses for various types of PACs, providing additional mechanistic insights into gene regulation in different AD phenotypes. Finally, we summarize our PACs into an AD-phenotypic single-cell atlas, and provide an open-source tool and web app for general use by the scientific community.

### Phenotype scoring of single cells

Our Phenotype Associated Single Cell encoder (PASCode) is a deep learning framework for scoring phenotype associations of cells (**Fig. 1a**). First, PASCode takes the population-level single-cell sequencing (e.g. snRNA-seq) dataset as input, with available donor-level phenotype information.

**Fig. 1:**
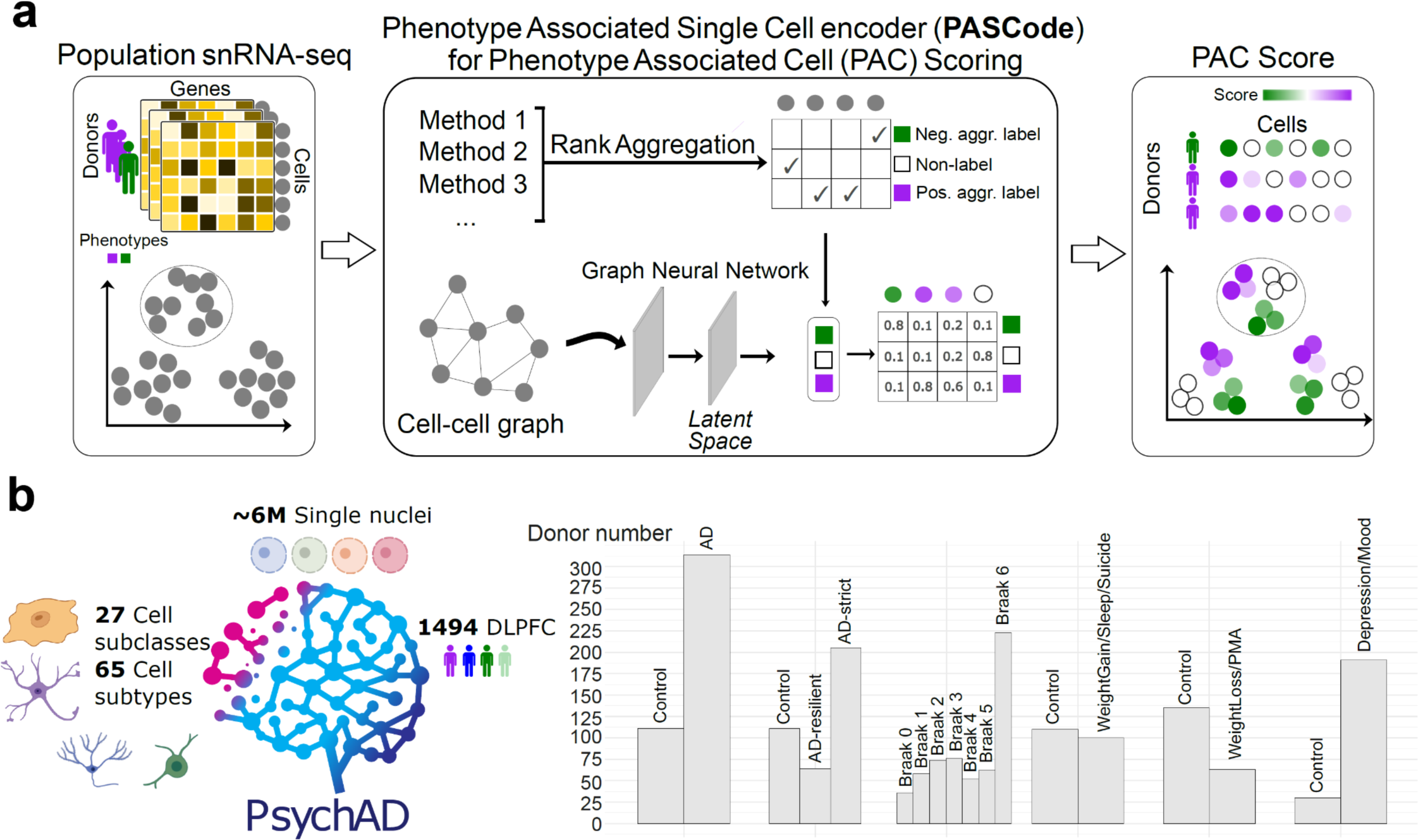
Phenotype scoring of single cells by Phenotype Associated Single Cell encoder (PASCode) with applications to the PsychAD dataset. a, PASCode takes a population-scale single-cell sequencing (e.g., snRNA-seq) dataset with phenotype labels as input. Multiple methods that identify phenotypic differentially abundant cells were applied to the dataset, followed by Rank Aggregation^14^ to generate robust ‘aggregated phenotype labels’ for all cells including Positive labels (Pos. aggr. label): cells associated with the positive condition of the phenotype; Negative labels (Neg. aggr. label): cells associated with the negative condition of the phenotype; Non-label: cells associated with neither condition. A graph attention network (GAT)^15^ model was trained to use the single-cell gene expression and the cell-cell similarity graph for predicting the ‘aggregated phenotype labels’. The trained GAT outputs a phenotype association score for each cell (PAC score), while providing a latent space that further subdivides cell types into subpopulations based on their cellular phenotypic association. More method details are in **Supplementary Note 1**. **b,** PASCode is applied to the snRNA-seq data from PsychAD consortium, which includes over 6 million single nuclei, 27 cell subclasses and 65 cell subtypes from 1,494 donors (dorsolateral prefrontal cortex, DLPFC). We specifically focused our analysis on 6 AD and neuropsychiatric symptoms (NPSs) phenotype contrasts (**Supplementary Note 1.3**) including Controls: AD, pathology-cognition (AD-strict and AD-resilient), AD progression (Braak stages), WeightGain/Sleep/Suicide, WeightLoss/PMA (weight loss and psychomotor agitation) and Depression/Mood.

Then, it applies a Robust Rank Aggregation algorithm^14^ to ensemble multiple differential abundance (DA) methods (**Supplementary Tables 1, 2**) and assigns aggregated phenotype labels to all cells (**Supplementary Notes 1.1.1, 1.1.2**). Given one phenotype contrast between a positive condition (e.g., AD) and a negative condition (e.g., Control), DA methods measure the per-cell importance for each condition^12^, followed by an aggregation that assigns ‘aggregated phenotype labels’ to each cell: ‘positive (negative) aggregated label’ assigned to cells consistently ranked as important for the positive (negative) condition across all the DA methods, while ‘non-label’ is applied to cells important for neither condition. For robust label assignment, we benchmarked several DA methods with synthetic data (**Supplementary Note 1.2.1** and **Supplementary** Figs. 1, 2, 4) and selected the most accurate ones for aggregation (**Supplementary Note 1.1.2**). Benchmark experiments validated that such an ensemble approach improved the accuracy of single-cell phenotype label assignment (**Supplementary** Figs. 1**, 2**).

Second, PASCode trains a graph attention network (GAT) model to classify the ‘aggregated phenotype labels’ of cells based on their gene expression (**Supplementary Note 1.1.3, Supplementary Note 1.2.2,** and **Supplementary** Fig. 3). After classification, the GAT model can assign a probability to quantify how one cell is associated with each aggregated label. We then defined the phenotype association score of the cell as its probability difference between positive and negative labels (i.e., phenotype associated cell score, or PAC score; **Supplementary Note 1.1.3**).

The pre-trained GAT model can be used to efficiently predict PAC scores for unseen cells^15^, while providing a latent space that further divides cell types into subpopulations based on cellular phenotypic associations. The GAT model can also be used to predict PAC scores for cells from donors with missing phenotypes, providing potential novel insights into phenotypes affecting these donors. With the predicted PAC scores, we can also prioritize important cell types for the phenotype using methods such as Random Forest^16^ and SHapley Additive exPlanations (SHAP)^17^; see **Methods**. Also, cells from each cell type can be subdivided into three subpopulations by cutting their PAC scores based on a predefined threshold (**Supplementary Note 1.1.4**): *PAC*^+^ cells associated with the positive condition (e.g., *AD*-*PAC*^+^for AD); *PAC*^−^ cells associated with the negative condition (e.g., *AD*-*PAC*^−^ for Control); *Non*-*PAC* cells not significantly associated with the phenotype of interest. Various analyses (e.g. differential gene expression, trajectory and gene regulatory network analysis) can be performed with these PACs to pinpoint the molecular mechanisms underlying the phenotype.

We applied PASCode to the population-level PsychAD consortium snRNA-seq data and scored the phenotype association of cells for multiple AD and NPS phenotypes. In total, PsychAD includes over 6 million cells (including 27 cell subclasses and 65 cell subtypes) from the DLPFC of 1,494 brain specimens (**Fig. 1b, PsychAD dataset in Supplementary Information**). We further preprocessed the data and performed feature selection and cell type annotation (**Methods**)^7^. Here, we focused on 584 donors with extensive AD phenotypic information (e.g., diagnosis, Braak stage, NPSs, **Extended Fig. 1a**) across various demographic conditions (e.g., ethnicity, sex, age), thereby covering a wide spectrum of AD-related phenotypes, including diagnosis, cognitively resilience, AD progression (Braak stages) and presence or absence of various NPSs (**Extended Fig. 1b, Supplementary Note 1.3**). Using the PAC scores of these phenotypes, we further identified cell-subpopulation-level associated genes and regulatory networks for AD and NPSs, as well as disease progression trajectories for the pathology-cognition contrast.

### Prioritizing cell subpopulations and genes in AD

Statistical tests to compare: (1) AD vs. Control AUCell score distributions at both PAC- and donor-level, and (2) the differences observed between AD and Control within PAC-level vs. donor-level, were conducted as described in **Supplementary Note 2.3** and tabulated in **Supplementary Tables 3, 4**. These tests suggest that PACs provide better separation between AD vs. Control compared to the donor-level. **g,** Statistical significance (adjusted p-values) of differential expression genes identified by comparing at Donor- vs. PAC-level (i.e., the difference between AD and Control captured by all the cells vs. by PACs) for microglia. Only genes with log2 fold change larger than 0.5 and adjusted p-value less than 0.05 were labeled. Genes highlighted in purple are those only found at the PAC-level, and in orange are those only found at the donor-level. Bolded genes are from three known AD associated microglia pathways (GO:001540, Amyloid-beta (Aβ) binding; GO:0048156, Tau protein binding; KEGG: hsa05010, Alzheimer disease-Homo sapiens) (**Supplementary Data 1**).

We first applied PASCode to calculate the AD association score (AD-PAC score) of cells using AD diagnosis information of the PsychAD donors: AD (n=314) and Control (n=111) (**Supplementary Note 1.3**). Equal numbers (n=100) of AD and Control donors with balanced male/female ratio were randomly selected as the training dataset (including 776,273 cells). As shown in **Fig. 2a** (top), cells from AD and controls were mixed in each cell subclass within the gene expression based UMAP plots. However, they became well separated in the trained PASCode latent space by both AD-PAC scores (**Fig. 2a**, bottom) and ‘aggregated phenotype labels’ for AD (**Supplementary** Fig. 6a). We then constructed three independent validation datasets, all from prefrontal cortex, to evaluate the pre- trained PASCode model (**Methods and Supplementary Note 2.1**): (1) PsychAD_heldout: subsampled 11 AD vs. 11 Control donors after excluding the 200 training dataset donors; (2) Seattle Alzheimer’s Disease Brain Cell Atlas (SEA-AD): 39 AD vs. 39 Control donors^19^; (3) ROSMAP: 24 AD vs. 24 Control donors^20^. These cohorts were input to the pre-trained PASCode model for predicting AD-PAC scores and compared them with the pre-calculated ‘aggregated phenotype labels’ as the ground truth (**Supplementary** Fig. 6b). Significant separation of AD-PAC scores was observed across cells of different ‘aggregated phenotype labels’: p < 10^-4^ using Wilcoxon rank-sum test (**Fig. 2b**), validating the capability of pre-trained PASCode models to predict PAC scores for unseen cells.

**Fig. 2:**
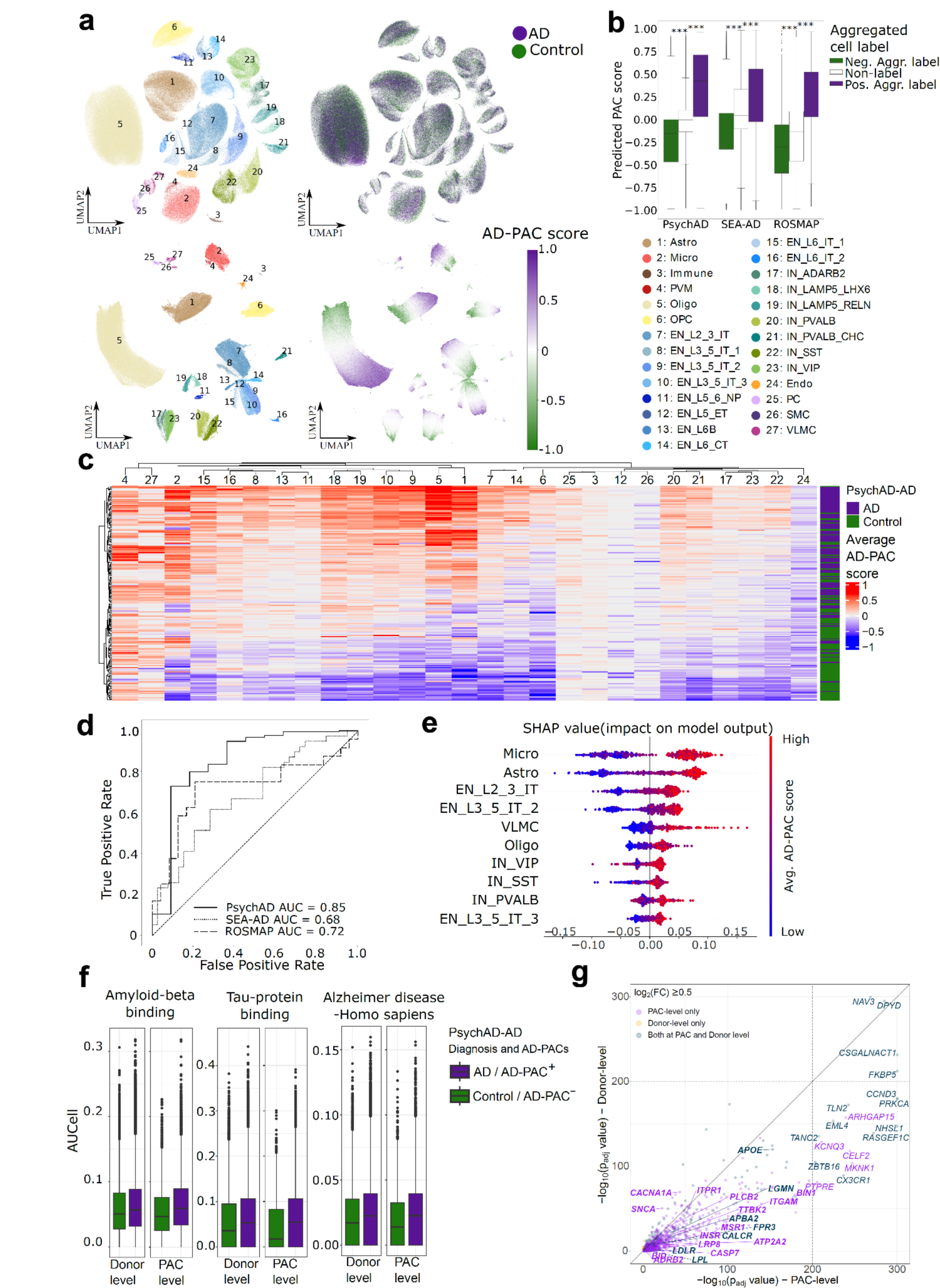
Alzheimer’s Disease associated cells to prioritize important cell types, genes and pathways for AD. a, Uniform manifold approximation and projection (UMAP)^18^ plots of gene expression (Top panel, Left: colored by cell subclasses; Right: colored by donor AD diagnosis) and PASCode latent space (Bottom panel, Left: colored by cell subclasses; Right: colored by AD-PAC score); **b,** The AD-PAC score predictions for three validation datasets: (1) PsychAD_heldout with AD (n=11) vs. Control (n=11), created by subsampling PsychAD AD vs. Control contrast after the training dataset donors were excluded, (2) SEA-AD^19^ with AD (n=39) and Control (n=39), and (3) ROSMAP^20^ with AD (n=24) and Control(n=24). The cells were grouped based on the pre-calculated aggregated labels: Purple - Positive Aggregated label (Pos. Agg. label) for AD. White - Non- label. Green - Negative Aggregated label (Neg. Agg. label) for Control.***: p < 10^-4^ based on Wilcoxon rank- sum test. **c,** Averaged AD-PAC scores of the 27 cell subclasses across PsychAD training dataset donors (n=100 AD vs. 100 Control). Rows: donors. Columns: cell subclasses. **d,** ROC curves for classifying AD vs. Controls in the three validation datasets described in **(b)**. The classification was performed by the Random Forest model using the average AD-PAC scores of 27 cell subclasses of donors in **(c).** Label transferring function ‘scanpy.tl.ingest’ in the Scanpy library^21^ was used to ensure SEA-AD and ROSMAP have the same cell type annotations with PsychAD (**Supplementary** Fig. 5). **e,** SHAP values for prioritizing AD associated cell subclasses using the Random Forest model in (**d**). Top 10 subclasses were displayed. **f,** AUCell scores on three known AD associated gene pathways of microglia at Donor-level (cells from AD vs. Control) versus PAC- level (*AD*-*PAC*^+^vs. *AD*-*PAC*^−^). Since genes in these pathways can be either upregulated or downregulated, we only included those upregulated genes as reported by Mathys *et al*.^20^ when calculating the AUCell scores.

Utilizing the AD-PAC scores, we further prioritized cell subclasses important for AD. To this end, for each donor in the training dataset (**Fig. 2c, row**), we averaged the PAC scores for each cell subclass to represent phenotype association on the cell subclass level (**Fig. 2c, column**). A Random Forest classifier that predicts AD versus Control was trained on the averaged PAC scores (**Fig. 2d**) and its SHAP values were utilized to prioritize cell subclasses important for AD (**Fig. 2e, Supplementary Data 1**). Among these, microglia were ranked the highest, followed by astrocytes, both of which have previously been associated with AD^22–24^. Although oligodendrocytes appear as the most prominent cell subclass with the greatest number of cells (**Extended Fig. 2a**), they were ranked relatively low, suggesting the PAC based cell type prioritization is not influenced by cell abundance^25^, unlike existing methods that utilize the number of differentially expressed genes^25,26^. We then grouped cells into PACs (i.e., *AD*-*PAC*^+^, *AD*-*PAC*^−^, *Non*-*PAC*) based on their PAC scores (**Supplementary Note 1.1.4**), and compared the proportion of *AD*-*PAC*^+/−^ within each cell subclass (**Extended Fig. 2a**). Neuronal subclasses, including both excitatory and inhibitory neurons, exhibited higher levels of *AD*-*PAC*^−^ compared to *AD*-*PAC*^+^, especially inhibitory neurons of the SST subclass (IN_SST) that showed ∼10 fold-change. This finding confirmed recent reports describing depletion of IN_SST in AD patients^27,28^. In contrast, non-neuronal subclasses, such as microglia, astrocytes, oligodendrocytes, and VLMC, showed an opposite trend, underscoring their different roles in AD.

Subsequent investigations focused on the expression of known AD genes within our predicted AD-PACs. AUCell was used to assess the enrichment of a given gene subset within each cell. Briefly, we first curated a list of upregulated genes in three known AD-related gene enrichment and pathway terms (e.g., tau-protein binding) for microglia (**Supplementary Note 2.2** and **Supplementary Data 1**) and investigated their enrichment in microglia at both the PAC-level (i.e., *AD*-*PAC*^+^vs. *AD*-*PAC*^−^) and donor-level (AD vs. Control cells based on donor phenotype labels) (**Fig. 2f**). Statistical tests using bootstrap subsampling and the Mann-Whitney U rank test (**Supplementary Note 2.3**) indicated that the AUCell scores are statistically more enriched in *AD*-*PAC*^+^than in *AD*-*PAC*^−^. Furthermore, the difference observed between AD and Control at the PAC-level is statistically more significant than that at the donor-level (**Supplementary Tables 3, 4**).

Additionally, we obtained a higher number of differentially expressed (DE) genes at the PAC- level than at the donor-level in microglia. Most DE genes identified at the donor-level overlapped with PAC-level DE genes (n=1,148 DE genes at the PAC-level vs. 220 DE genes at the donor-level, with 198 overlapping), although the statistical significance of donor-level DE genes was much lower (**Fig. 2g**). Notably, many of the PAC-level specific DE genes were related to known AD microglial functions, e.g., tau-protein binding and Aβ binding (**Fig. 2g, Supplementary Data 1**). We also extended the DE analysis to the other five top prioritized cell subclasses: astrocytes, VLMCs, two intratelencephalic (IT) excitatory neuronal cell types and oligodendrocytes, allowing us to achieve greater statistical significance for DE genes at the PAC-level compared to the donor-level across all these subclasses (**Extended Fig. 2b and Supplementary** Fig. 7a-d).

Lastly, we examined how PAC-level DE genes are enriched in the validation datasets. *AD*-*PAC*^+^upregulated genes were curated for each cell subclass (**Supplementary Data 1**) and used as gene expression signatures (*AD*-*PAC*^+^_*gene*_*set*). We calculated the AUCell scores using *AD*-*PAC*^+^_*gene*_*set* for the entire PsychAD cohort (i.e., both training and test datasets) and visualized their distributions at the donor-level (cells from AD vs. Control) versus PAC-level (*AD*-*PAC*^+^ vs. *AD*-*PAC*^−^) (**Extended Fig. 2c**). The differences within PACs were much more significant than those at the donor-level **(Supplementary Tables 5, 6, Supplementary Notes 2.2, 2.3**). We also extended our analysis to two independent datasets for validation (SEA-AD and ROSMAP) (**Extended Fig. 2d, e**).

Together, these results provided two forms of validation: (1) PAC-level DE genes were consistently enriched within the AD donors of independent datasets at both the PAC- and donor-level, indicating that these DE genes are indeed AD related; and (2) the AUCell score differences at the PAC-level were more significant than those at the donor-level in the independent datasets, indicating the utility of using PACs (compared to using all cells) for disease biomarker identification. Additionally, we employed Gene Set Variation Analysis (GSVA, **Supplementary Note 2.2**) to validate the *AD*-*PAC*^+^_*gene*_*set* within another independent AD bulk RNA-seq dataset^29^. We found that the enrichment scores significantly increased along the AD Braak stage (**Extended Fig. 2f-h, Supplementary** Fig. 7e-g), especially for those prominent cell subclasses such as microglia, astrocytes and oligodendrocytes, suggesting potential roles for genes within the *AD*-*PAC*^+^_*gene*_*set* in AD progression.

### Gene expression dynamics in AD progression and resilience

Alzheimer’s disease is characterized by progressive accumulation of neuropathological lesions and cognitive decline^31^. To describe the severity of AD progression, metrics such as Braak stage, CERAD score, and CDR score have been developed^32^. However, these clinical measurements are generally heterogeneous and hysteretic, and, although studies focusing on evaluating neuropathological pseudotime have been conducted, they have primarily used bulk RNA-seq or imaging based approaches^33,34^. Here, we sought to understand AD progression at single-cell level by scoring cells associated with Braak stage (i.e., *AD*-*progression*-*PAC*). To do this, we first isolated 30 donors from Braak stage 0, representing control or early-Braak-stage donors, and 30 donors from Braak stage 6, representing late-Braak-stage donors (**Fig. 3a**, right), as training data for PASCode (**Fig. 3a**, left). The trained PASCode model allows scoring AD progression association for cells from donors of the intermediate Braak stages (i.e., Braak stages 1-5). Similar to **Fig. 2** analysis, we trained a Random Forest classifier on the averaged *AD*-*progression*-*PAC* score across cell subclasses. The classifier output indicates the similarity of the donors’ PAC scores compared with the training donors (i.e., Braak stages 0,6), and can be taken as ‘AD progression stage time’ that mimics the pseudo- progression of AD (**Methods**). As shown in **Fig. 3b**, the predicted ‘AD progression stage time’ significantly correlated with Braak stages (p < 10^-4^ based on Jonckheere-Terpstra test), suggesting that the neurofibrillary tangles changes observed with AD progression were captured by the ‘AD progression stage time’. Although with much fewer donors with AD (n=30) vs. Control (n=30) than **Fig. 2** analysis with AD (n=100) vs. Control (n=100), cell subclass prioritization resulted in highly consistent rankings (**Fig. 2e, Supplementary** Fig. 8b), indicating the robustness of PASCode.

**Fig 3.**
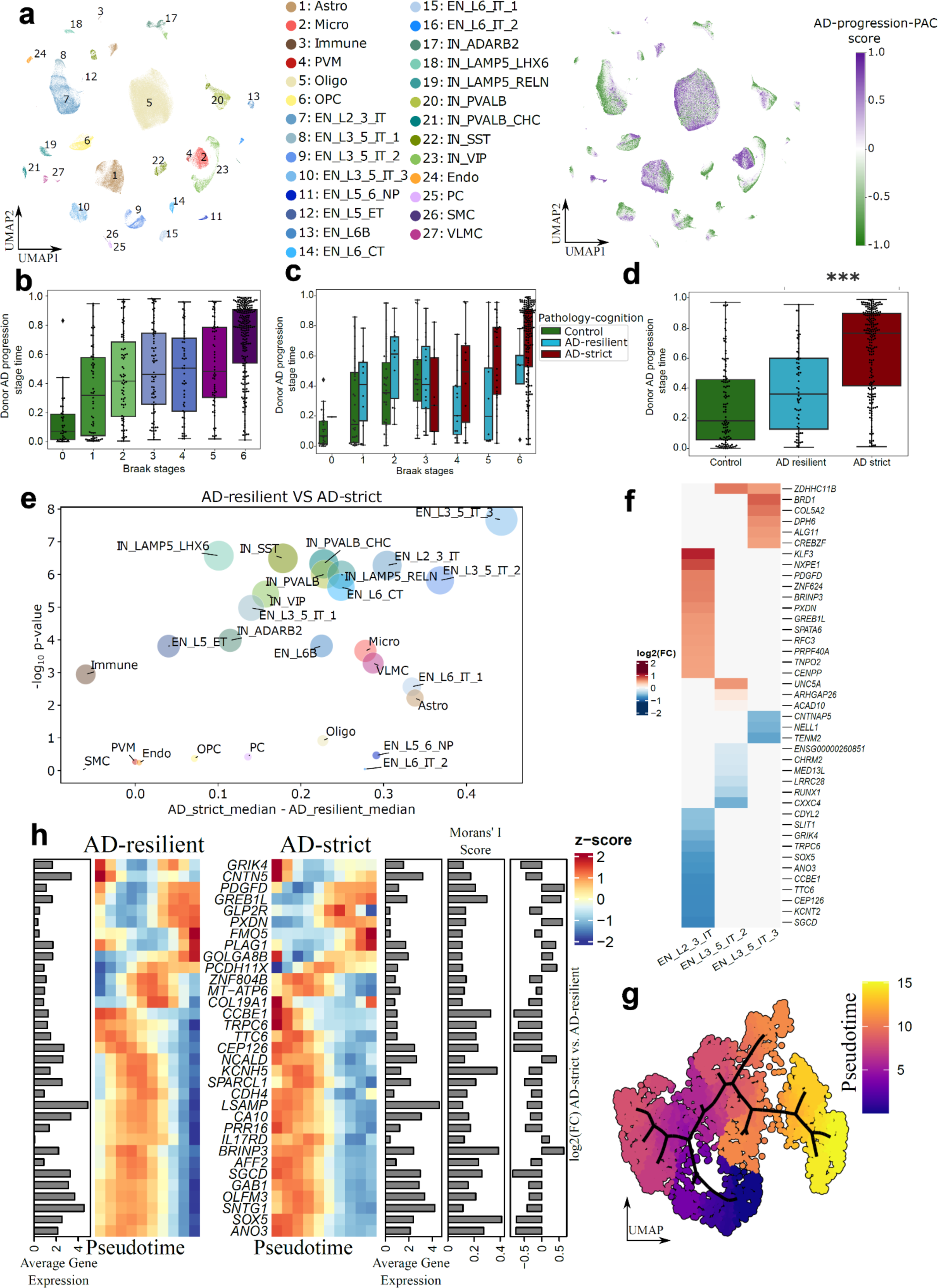
Phenotype associated cells capture the cell-subpopulation-level gene expression dynamics in Alzheimer’s disease progression and resilience. a, PASCode latent space based UMAP plots depicting cell subclasses (left) and *AD*-*progression*-*PAC* scores (right). Only the training dataset (n=30 donors each from Braak stages 0 and 6) is included for visualization. **b,** ‘AD progression stage time’ (defined based on *AD*-*progression*-*PAC*, see **Methods**) distribution of donors across all Braak stages. **c,** ‘AD progression stage time’ distribution across the pathology-cognition contrast (**Supplementary Note 1.3**, Green: Control, Cyan: AD-resilient, Maroon: AD-strict) within each Braak stage. **d,** ‘AD progression stage time’ distribution across pathology-cognition contrast for all donors. ***: p < 1.22×10^-10^ based on one-sided student’s t test. **e,** Bubble plot to show the difference of SHAP values across AD-resilient and AD-strict donors. Both Y-axis and bubble size represent minus log10 Wilcoxon rank-sum test p-values comparing the SHAP values. To exclude the stochastic effects, we trained 100 Random Forest models with different random seeds, and averaged their SHAP values for comparison. X-axis is the difference in the median values of the averaged *AD*-*progression*-*PAC* score between AD-resilient and AD-strict donors. **f,** DE genes across AD-resilient vs. AD-strict donors for EN_L3_5_IT_3, EN_L3_5_IT_2, and EN_L2_3_IT. DE gene analyses were conducted using *AD*-*progression*-*PAC*^+^ cells (i.e., cells associated with late-Braak-stages) with sex and Braak stage groups as covariates. Only genes that satisfy adjusted p-value ≤ 0.05 and |log2(fold change)| ≥ 0.5 are depicted in the heatmap (number of total DE genes without the fold change threshold, EN_L2_3_IT: 213, EN_L3_5_IT_2: 10, EN_L3_5_IT_3: 20). **g,** AD progression trajectory of EN_L2_3_IT based on the top 5000 highly variable genes in *AD*-*progression*-*PAC*^+^. The cells are colored based on inferred pseudotime (**Methods**). **h,** Pseudo-temporal variation of AD progression genes that are differentially expressed across AD-resilient (left) and AD-strict (right) PACs. Genes were selected by overlapping AD progression genes (trajectory driver genes with Moran’s I score ≥ 0.1 and q-value < 0.01) and DE genes in **Supplementary Data 2** corresponding to panel **(f)**. Bar plots represent the Moran’s I score, average gene expression, and the log2(FC) values obtained from AD-strict vs. AD-resilient DE gene analysis. The heatmap depicts the standardized gene expression variation along pseudotime.

Interestingly, the ‘AD progression stage time’ distributed differently across AD-resilient (potential AD resilience) and AD-strict donors of the pathology-cognition contrast (**Supplementary Note 1.3**), especially within the late Braak stages (p < 0.86, 0.052, 0.021, 0.041 by one-sided student’s t test for Braak stages 3,4,5,6 respectively. **Fig. 3c-d, Supplementary** Fig. 8a). This suggests that AD donors with preserved cognition may have distinct molecular mechanisms that differentiate them from their cognitively compromised counterparts, even at the same stage of neurofibrillary tangle pathology (Braak). To identify the cell subclasses that most differ across AD-resilient and AD-strict, we conducted statistical tests using the Random Forest SHAP values (**Fig. 3e, Supplementary Data 2**) and showed that the majority of neuronal cell subclasses (EN and IN) were significantly differentiated, indicative of an important role for neuronal cells in resilience to AD. Among them, IT ENs (EN_L3_5_IT_3, EN_L3_5_IT_2, EN_L2_3_IT) were ranked as the most differentiated, followed by several IN subclasses (IN_PVALB_CHC, IN_SST, IN_LAMP5_RELN, IN_LAMP5_LHX6, IN_VIP). These observations are supported by recent studies that found IT EN^28^ and some IN cell subclasses^27,28^ like IN_SST, IN_LAMP5_RELN are associated with cognitive resilience in AD. Also, astrocytes and microglia were among the most differential cell subclasses identified in our analysis.

Notably, Mathys *et al*. also identified astrocytes to associate with cognitive resilience in a recent study^35^. Thus, these results suggest the ‘AD progression stage time’ based on *AD*-*progression*-*PAC* scores capture the underlying molecular mechanisms of resilience to AD progression.

To identify gene expression differences underlying AD progression between AD-strict and AD-resilient donors, we conducted DE analysis comparing *AD*-*progression*-*PAC*^+/−^ within the top 3 IT cell subclasses: EN_L2_3_IT, EN_L3_5_IT_2, and EN_L3_5_IT_3 (**Fig. 3f** and **Supplementary Data 2**). The DE analysis was conducted using MAST^36^ with sex and Braak stage as covariates. For instance, the *BRD1* gene is highly enriched in AD-strict within EN_L3_5_IT_3 subclass and was reported to contribute to cognitive impairment in mice^37^. *TENM2* (Teneurin transmembrane protein 2) is among the genes downregulated in AD-strict EN_L3_5_IT_3 cells and is associated with spine genesis and synapse maturation^38^. *UNC5A* is upregulated in AD-strict EN_L3_5_IT_2 cells, and animal models suggest it contributes to AD via activating death-associated protein kinase 1 (*DAPK1*)^39^. Logan *et al*.^40^ have demonstrated that *RUNX1* promotes neuronal differentiation to facilitate repair of neuronal function following injury. Being enriched in AD-resilient cells could potentially be an indication of the *RUNX1* gene’s role in protecting cognitive function in AD patients.

Schizophrenia and ASD-associated gene^41^ *CXXC4*, encoding a transcription factor antagonizing Wnt signaling, was also found to be enriched in AD-resilient EN_L3_5_IT_3. The gene *KCNT2*, encoding KNa1.2 a chloride-activated potassium channel, is among the most down-regulated genes in AD-strict EN_L2_3_IT. Loss of function of KNa1.2 in mice leads to motor deficits and enhances seizure susceptibility^42^, and could be one of the critical genes that differentiate impaired and preserved cognition, which has been associated with AD^43^.

To further investigate genes related to AD progression, we inferred a single-cell trajectory for EN_L2_3_IT, the most affected subclass in AD-resilient compared to AD-strict donors, using the top 5000 highly variable genes within *AD*-*progression*-*PAC*^+^(**Fig. 3g**, and **Extended Fig. 3a,b**).

Differential progression along the pseudotime (**Extended Fig. 3c**) shows variation across AD-resilient vs. AD-strict donors (left) and Braak stages (right). As expected, Early-Braak (i.e., Braak stages 0-2) are clustered at earlier times, and the severity of AD increases with the inferred pseudotime.

Similarly, most AD-resilient cells spread in early-mid pseudotime, while AD-strict cells spread more in mid-late pseudotime. Gene Ontology (GO) functional enrichment of the AD progression driver genes (**Supplementary Data 2**) (Moran’s I score > 0.1 and q.value < 0.01) displayed many synapse-related terms (glutamatergic synapse), dendrite-related terms (dendritic tree^44^) and cellular organization and survival (catenin complex^45^, calcium−dependent cell−cell adhesion via plasma membrane cell adhesion molecules^46,47^. Among them, the glutamatergic synapse has previously been reported as a factor leading to calcineurin that contributes to the phosphorylation of tau and ubiquitin proteins^48–50^.

To investigate gene expression dynamics across AD progression and resilience, we overlapped AD-progression driver genes with DE genes across AD-resilient and AD-strict (**Fig. 3f, Supplementary Data 2**). **Fig. 3h** depicts the pseudo-temporal variation of these genes in AD-resilient (left) and AD-strict (right) groups. *BRINP3*, *KCNH5, TRPC6* and *SOX5* were found to be highly variable along the AD progression with a Moran’s I score >0.3. *KCNH5* is upregulated in our AD-resilient cells, supporting a previous observation in neuronal cells of mice that are resistant to AD^51^.

*TRPC6*, encoding a receptor to activate the calcium channel, exhibited a more dramatic change in AD-strict than AD-resilient. *TRPC6* dysfunction has been associated with cognitive deficits in AD^52^ and activation of *TRPC6* has been used as a potential target for antidepressants^53^. *SOX5* is a known candidate gene for late-onset AD that plays an important role in neuronal development^54^. The increasing activity of *SOX5* along AD progression correlates with Braak stages in both AD-strict and AD-resilient cells, suggesting that *SOX5* may be involved in regenerative responses. Interestingly, the layer L2/3 glutamatergic neuronal marker, *CCBE1*^55^, also showed dynamic variation along AD progression and was upregulated in AD-resilient cells.

One common observation from both the donor-level ‘AD progression stage time’ (**Fig. 3c**) and PAC-level pseudotime analysis (**Extended Fig. 3c**) is that, while AD-resilient donors/cells spread across mid-late stages of AD progression, AD-strict donors/cells mainly clustered in the later stages. AD-strict donors correspond to more severe progression than AD-resilient donors across later Braak stages (**Fig. 3c**). Thus, comparison across AD-strict vs. AD-resilient would be more accurate if similar disease stage donors are considered. Therefore, we estimated the cell subtype level rankings for donors with Braak stages above 3 (**Extended Fig. 3d**). Similar to **Fig. 3e**, neuronal cell subtypes show the highest significance of SHAP value differences across AD-resilient and AD-strict. Among non-neuronal cells, astrocyte subtype Astro_PLSCR1 shows the highest significance, followed by Astro_ADAMTSL3 and Immune_B subtypes. Notably, astrocytes subtypes show a considerable variation in SHAP value differences, highlighting the importance of conducting analysis in subpopulations of the canonical cell types. As such, we focused our analysis on the four astrocyte subtypes using *AD*-*progression*-*PAC*^+^and examined the gene expression in neurotoxic (A1) and neuroprotective (A2) reactive astrocytes^56^ (**Extended Fig. 3e**). Among the neurotoxic (A1) markers, *FKBP5* appears to be expressed in all astrocyte subtypes across both AD-resilient and AD-strict donors. However, pairwise comparison (**Extended Fig. 3f, top**) indicates Astro_ADAMTSL3 and Astro_PLSCR1 subtypes show increased expression in AD-strict cells. High levels of *FKBP5* have been associated with cognitive deficits in animal models^57^. On the other hand, the neuroprotective markers, *EMP1* and *CD109* are expressed mainly in Astro_PLSCR1 and Astro_ADAMTSL3, respectively. *EMP1* expression in Astro_PLSCR1 depicts a significant upregulation in AD-resilient cells (**Extended Fig. 3f**). *EMP1* is a membrane protein involved in cell survival of cancer cells^58^ and its function in astrocytes, or in the brain, is largely unknown. Reduced *EMP1* expression levels have been found in a population of patients with major depressive disorder^59^. This differential expression of neuroprotective and neurotoxic markers across AD-resilient and AD-strict indicates that the Astro_PLSCR1 subtype may contribute to neuronal damage resistance in AD-resilient individuals.

### Depression associated cells in AD

Clinical studies suggest links between depression symptoms and AD^60^. We applied PASCode to score depression (Depression/Mood contrast, **Supplementary Note 1.3**) associated cells by comparing AD donors (AD-resilient donors excluded) with (n=100) and without depression (n=110) (**Fig. 4a**), thus excluding AD-specific signals. Interestingly, we observed *Depression*-*PAC*^+/−^distributed across all the cell subclasses (**Fig. 4b, 4c**), indicating a global alteration of cell states as a result of depression in AD. In particular, astrocytes and oligodendrocytes were prioritized as the top two cell subclasses associated with depression (**Fig. 4d**,). Furthermore, zooming into cell subtype level also showed Astro_WIF1 (wnt inhibitory factor 1-expressing), Oligo_OPALIN (Oligodendrocytic Myelin Paranodal And Inner Loop Protein), Astro_GRIA1 (astrocytes expressing *AMPA* receptor *GluA1* can modulate neural transmission of neurons), and Astro_ADAMTSL3 (*ADAMTS* Like 3) as the highest prioritized cell subtypes (**Fig. 4e, Supplementary Data 3**). Astro_WIF1, a major component of protoplasmic astrocytes^61^, contributes to regulating synaptic transmission and supporting metabolically neurons^62^, while Astro_ADAMTSL3 is a part of fibrous astrocytes^61^.

**Fig. 4.**
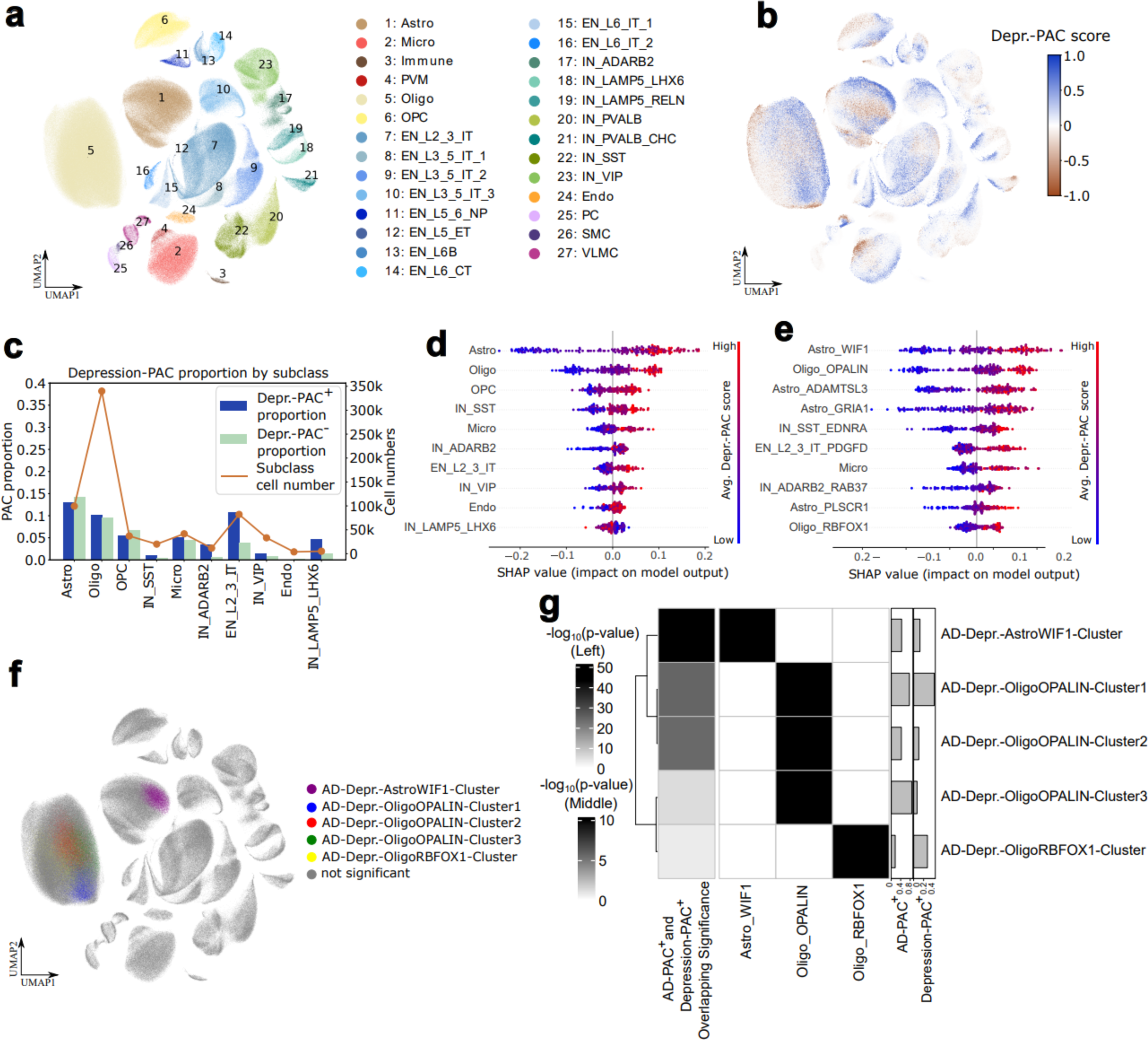
Depression associated cells to pinpoint cell subpopulations important for depression within AD. a-b, UMAP visualization of cells from AD donors with available depression information. Cells are colored by cell subclasses (**a**) and Depression-PAC score (**b**). **c,** Proportion of *Depression*-*PAC*^+/−^(i.e., cells associated with depression (*Depression*-*PAC*^+^) or control (*Depression*-*PAC*^−^)) within each of the top 10 prioritized depression associated cell subclasses shown in (**d**). Orange curve represents the number of cells within each cell subclass as labeled on the right y-axis. **d-e,** SHAP values for prioritizing depression associate cell types at subclass-level (**d**) and subtype-level **(e)**; top 10 prioritized cell types are shown. **f,** Cell clusters that are significantly overlapped between *AD*-*PAC*^+^and *Depression*-*PAC*^+^. Only clusters with more than 1000 cells, a percentage of *PAC*^+^greater than 5%, and p-values less than 0.01 (with hypergeometric test) were considered. **g,** Detailed information of significant overlapped cell clusters in (**f**). Left: log10 transformed hypergeometric p-values of the overlapping significance between *AD*-*PAC*^+^and *Depression*-*PAC*^+^within each cluster. Middle: log10 transformed hypergeometric p-values of the overlapping significance of each cell cluster and cell subclass. Right: Fraction of *AD*-*PAC*^+^and *Depression*-*PAC*^+^within each cluster.

Together, these observations suggested astrocytes and oligodendrocytes likely play important roles in depression symptoms within AD donors. DE gene analysis between *Depression*-*PAC*^+/−^(**Supplementary Data 3**) revealed activation of inflammation within depression associated astrocytes (**Extended Fig. 4a**), consistent with previous studies^63–66^. We, therefore, screened for common inflammation markers and found *IL18* was significantly upregulated in *Depression*-*PAC*^+^compared to *Depression*-*PAC*^−^ astrocytes (**Extended Fig. 4b, c**). Besides astrocytes, *IL18* was also upregulated by *PAC*^+^ in other cell types (**Extended Fig. 4c**), substantiating the importance of this cytokine in depression^67–71^.

Further, cells associated with both AD and depression (overlap of *AD*-*PAC*^+^ with *Depression*-*PAC*^+^, hereafter referred as AD-Depression-PACs) may represent the mechanistic interplay between these phenotypes. To determine cell subpopulations significantly enriched with AD-Depression-PACs, we first clustered cells from all the donors based on their gene expression similarity. Then, for each cluster, we used hypergeometric tests to evaluate the overlapping significance between *AD*-*PAC*^+^ and *Depression*-*PAC*^+^(**Fig. 4f, Supplementary** Figs. 9**, 10**). Cell subpopulations significantly enriched with AD-Depression-PACs belong to Astro_WIF1 and Oligo_OPALIN subtypes (**Fig. 4g**), which were also prioritized as the most important subtypes for depression (**Fig. 4e**). Unlike for depression, only the protoplasmic astrocyte subpopulation (i.e., Astro_WIF1) was identified as contributing to both AD and depression. WIF1 (wnt inhibitory factor 1-expressing) is an inhibitory factor of the Wnt/β-catenin signaling pathway^72^, activation of which results in neuroprotection against Aβ^73^ and depression^74^. Finally, we analyzed genes significantly upregulated (n=271) in AD- Depression-AstroWIF1-cluster compared to the rest of the Astro_WIF1 cells (**Supplementary Data 3**). GO enrichment analysis found that they are enriched in Endoplasmic Reticulum stress pathways (**Extended Fig. 4d**), suggesting that protein misfolding may contribute to both AD^75^ and depression^76^. In addition, the enrichment of AD-Depression-PACs within Oligo_OPALIN, a subtype important for oligodendrocyte differentiation^77^, suggests that neuro-regeneration may also play a role in AD associated depression.

### Astrocyte gene networks in depression

Given the capability of PASCode to detect phenotype associated cells (PACs), we were next interested in a systems-level analysis to illuminate processes that drive gene regulatory mechanisms manifesting in these PACs. The regulation of gene expression often involves the interplay between several transcription factors (TFs). Regulatory modules, or sets of co-regulated genes, tend to cluster in networks and are often functionally related^79,80^. At the single-cell level, network modules can pinpoint pathways and genes that are dysregulated in disease phenotypes.

Given that astrocytes are implicated as the most distinct cell type across *Depression*-*PAC*^−^and *Depression*-*PAC*^+^(**Fig. 4**), we focused on examining gene network modules underpinning depression. To achieve this, we constructed gene co-regulation networks by connecting genes regulated by similar TFs (**Methods** and **Supplementary Note 3**). Subsequent clustering of these networks unveiled a robust modular organization within *Depression*-*PAC*^−^ and *Depression*-*PAC*^+^astrocytes (i.e., gene co-regulation modules; **Supplementary Data 4**). These modules are enriched with expected pathways and disease ontology terms, providing a comprehensive representation of astrocyte subtype biology (**Fig. 5 a-c**, **Supplementary** Fig. 11**, Supplementary Data 4**). For instance, Module 2 (M2) and Module 9 (M9) in depression associated astrocytes are enriched with genes related to neuroinflammation and glutamatergic signaling, and cholesterol biosynthesis, respectively (**Fig. 5a**). Genes within M2 and M9 are relatively more highly expressed in PLSCR1 and GRIA1 subtypes, respectively (**Fig. 5b**), reflecting the generalized role of these subtypes in overall astrocyte function. Interestingly, disease ontology enrichment analysis suggests the presence of depression-related modules within *Depression*-*PAC*^+^cells and the absence of AD-related modules within these cells (**Fig. 5c**). This indicates a good separation of depression associated cells and related pathways within astrocytes in AD.

**Fig. 5.**
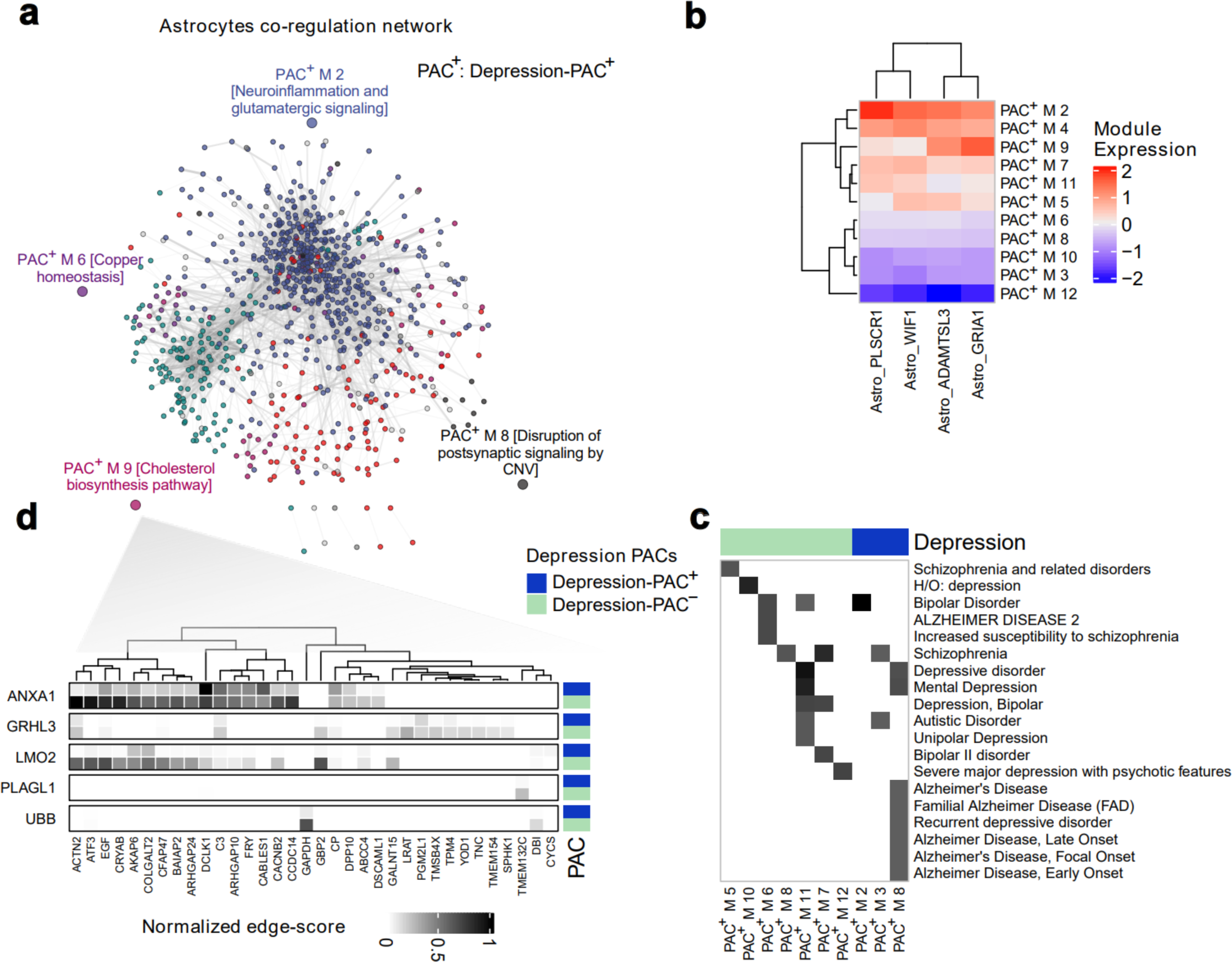
Gene network modules in depression associated astrocytes. **a**, The gene co-regulation network for *Depression*-*PAC*^+^ within astrocytes with individual genes represented as circles connected based on the overlap between their predicted regulators (i.e., gene co-regulation network modules; **Methods** and **Supplementary Note 3**). Genes are colored based on module membership. **b**, Average expression of genes within each module is shown across different astrocyte subtypes. **c**, The network modules identified in (**a**) were subjected to enrichment analysis against the DisGeneNet database^81^. Disease terms with statistical significance (FDR corrected hypergeometric test p-value < 0.1) are colored black in the heatmap grids. **d**, Top rewired TFs (y-axis) and their differential usage of *Depression*-*PAC*^+^ M9 geneset (x-axis) in *Depression*-*PAC*^+^ versus *Depression*-*PAC*^−^ astrocytes networks in (**a**). The heatmaps matrix is colored along a black gradient indicating normalized edge importance scores with darker colors indicating a stronger relationship between the TF and its target. The top row of each TF split represents *Depression*-*PAC*^−^ and the bottom row represents *Depression*-*PAC*^+^.

We further probed co-regulation modules to identify interesting TFs that rewire between *Depression*-*PAC*^−^ and *Depression*-*PAC*^+^ cells and found several instances of differential usage of target genes by TFs. For instance, the geneset in M9 exhibits considerable rewiring of its regulators in the *Depression*-*PAC*^+^. The TF, *ANXA1* (Annexin A1), known for its neuroprotective effects^82^, appears to exhibit weaker connection to its predicted targets in the M9 geneset in *Depression*-*PAC*^+^compared to *Depression*-*PAC*^−^ astrocytes (**Fig. 5d**). *GRHL3* is primarily known as a regulator of neurodevelopment and its deficiency has been shown to lead to deficits in brain and spinal cord development^83^. Our analysis also implicates *LMO2* as another example with differential targets in *Depression*-*PAC*^−^ and *Depression*-*PAC*^+^ cells. *LMO2* has been reported as a part of a transcriptional complex important in neurogenesis in chick embryos^84^. This suggests that GRHL3 and LMO2 may have altered interactions with their target genes in the context of *Depression*-*PAC*^+^, leading to changes in astrocyte function and potentially impacting neuroinflammatory processes and neuronal support. Understanding these changes could be crucial for identifying new molecular targets for therapeutic intervention in depression.

### Phenotypic single-cell atlas for AD and NPS

In addition, we scored cells association with other NPS phenotypes (**Supplementary Note 1.3**) in the PsychAD cohort and summarized the results as a phenotypic single-cell atlas. We anticipate that this atlas will serve as a valuable resource of gene expression and regulation mechanisms underlying AD and related phenotypes, and will facilitate more extensive studies of AD. Overall, this atlas encompasses ∼2.3 million single cells with phenotype association scores, ∼1.5 million PACs (**Supplementary Data 5**), and differentially expressed (DE) genes identified from the PACs. To understand the relative functional importance of the cell types in each phenotype, we prioritized the 27 subclasses based on their averaged PAC scores (**Supplementary Data 6**). For example, microglia and astrocytes have been prioritized as crucial for differentiating AD donors from controls (**Fig. 6b**). More complicated analyses can be conducted with PACs, such as the astrocytes subpopulations we found to be important for pathology-cognition (**Extended Fig. 3**) and AD- Depression interplays (**Fig. 4f-g**). We also summarized and compared the DE genes in AD with all the NPSs to gain insights into their possible relationships. For example, within oligodendrocytes, we identified 11 intersected upregulated genes between AD with WeightGain/Sleep/Suicide, 9 with WeightLoss/PMA (weight loss and psychomotor agitation), and 3 with Depression/Mood (**Fig. 6c**).

**Fig. 6.**
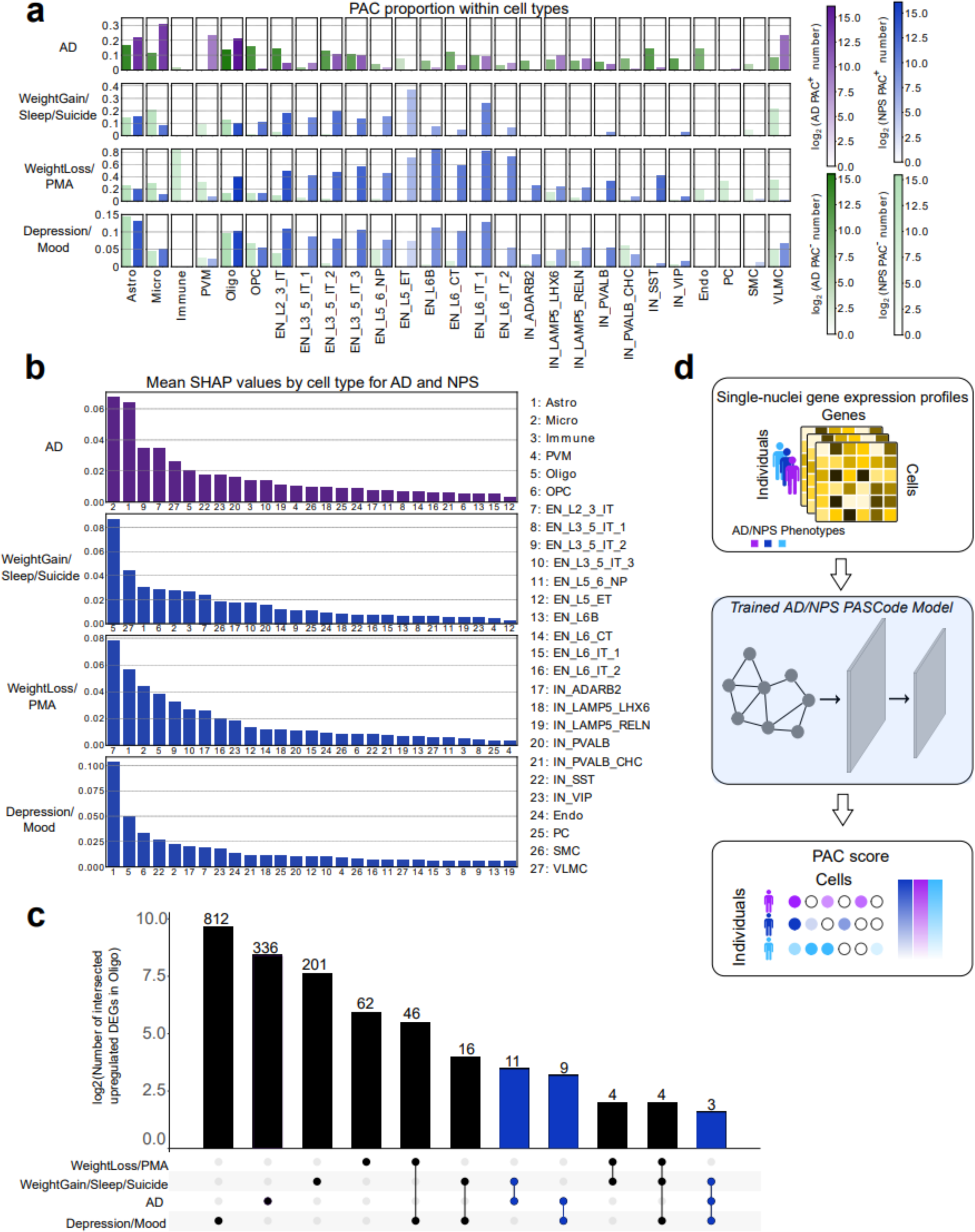
Phenotype scoring of single cells and differentially expressed genes atlas for AD, WeightGain/Sleep/Suicide, WeightLoss/PMA, and Depression/Mood in PsychAD. a, Proportion of *PAC*^+/−^associated with AD, WeightGain/Sleep/Suicide, WeightLoss/PMA (weight loss and psychomotor agitation) and Depression/Mood across subclasses. **b,** Cell type prioritization based on the mean absolute SHAP values across the four phenotypes. **c,** Number of overlapped upregulated genes between AD and NPS within oligodendrocytes. Bars are log2 transformed, and the exact numbers of overlapped genes are shown on top of each bar, such as AD and WeightGain/Sleep/Suicide (n=11), AD and WeightLoss/PMA (n=9), AD and Depression/Mood (n=3). **d**, PASCode models pre-trained on PsychAD facilitate assigning PAC scores for future snRNA-seq datasets.

These overlapped genes exhibit similar expression patterns across the relevant phenotypes, suggesting their potential roles in multiple phenotypes. A comprehensive summary of PAC-based DE genes for all the cell subclasses is provided in **Supplementary Data 7**. To facilitate the use of this atlas, we provide a web application for exploring these identified PACs (**Methods**). The pre-trained PASCode models are also provided to enable phenotype scoring of other single-cell datasets (**Methods**, **Fig. 6d**).

## Discussion

In this work, we used PASCode to score phenotype association (PAC score) of cells with various Alzheimer’s disease phenotypes, including progression, potential resilience, and associated neuropsychiatric symptoms. Our results suggest that PAC scores have improved the discovery of disease associated cell types, genes and pathways in AD. For instance, analysis based on AD-progression-PAC scores prioritized reactive astrocytes and several neuronal subpopulations as important in AD resilience during disease progression. Specific astrocyte subpopulations with upregulated inflammation pathways were also found to play an important role in AD associated depression. Network analysis further demonstrated that a systems-level approach of PAC analysis can recapitulate gene regulatory mechanisms and implicate cell subpopulation gene modules in specific phenotypes such as depression. However, this work only covers limited phenotypes within PsychAD. Future work will aim to summarize PACs for a broader spectrum of phenotypes. Using the atlas, researchers can directly explore genes or cells of interest within or across phenotypes. More complicated analysis of this atlas, such as the PAC mediated prediction of phenotype related gene regulatory networks, can also be performed.

PASCode can also be generalized to find associated single cells for other disease types and clinical phenotypes using population-scale single-cell data. The pre-trained PASCode model can also predict PACs in single-cell datasets from new donors, even without phenotype information. A recently published approach^85^ that can accomplish a similar purpose uses scArches ^86^ to map the new cells into a common latent space and then uses Milo^87^ to determine PACs within the latent space. Unlike PASCode, scArches and Milo must be re-run whenever new donors are available. The authors found that involving a healthy reference atlas for learning the latent space could improve PAC identification accuracy. Thus, in the future, PASCode can easily incorporate a healthy reference atlas as negative training samples for model training, or through transfer learning architectures^88^. We can further extend our analysis to other brain regions (e.g., middle temporal gyrus within SEA-AD^19^), resulting in an AD phenotypic atlas for the whole human brain. Moreover, we can integrate emerging single-cell multimodal data, such as scATAC-seq or spatial data, leading to a more comprehensive functional genomic atlas of disease associated cells.

## Methods

### PsychAD snRNA-seq data, data preprocessing, feature selection and cell type annotation

The PsychAD data description is available in “PsychAD dataset” (**See Supplementary Information**). We preprocessed PsychAD snRNA-seq data as described in Lee *et al*.^7^. Briefly, batch correction was performed with Harmony^89^ on the log2CPM gene expression values. The 50 Harmony Principal Components were used for constructing the KNN graph as input for the DA tools and GAT model. We identified the top 5,000 highly variable genes (HVGs) using the ‘highly_variable_genes()’ function in Scanpy (v.1.9.3)^21^ and focused on the 3,401 protein coding genes among them as features for PAC identification. To obtain the list of HVGs, each gene’s expression values across all cells were transformed to a standardized score with zero mean and unit standard deviation. Then, normalized variance of each gene was calculated from the transformed scores and genes exhibiting the largest variance were selected as HVGs. As described by Lee *et al*.^7^, 27 cell subclasses and 65 cell subtypes were identified based on their marker genes. We analyzed PACs distribution across these annotated cell types in this work.

### Phenotypes and neuropsychiatric symptoms in Alzheimer’s disease

This work predicted and analyzed phenotype associated cells (PACs) obtained for multiple phenotypes and neuropsychiatric symptoms (NPSs) in AD. Specifically, PsychAD leveraged a set of 19 NPSs commonly associated with AD and related dementias, which significantly impact daily functioning and quality of life^4^. Thus, to decipher the possible relationship across these phenotypes, we focused our analysis within the cohort (Mount Sinai NIH Neurobiobank, MSSM)^7^ containing clinical information of both AD phenotypes and NPSs. Out of the 1,042 MSSM donors, we selected 584 donors with extensive phenotypic information (e.g., diagnosis, Braak stage, NPSs; **Extended Fig. 1a**). Hierarchical clustering analysis also found that the 19 NPSs tend to form three distinct clusters, generally consistent with known associations^7^. Therefore, considering the sample sizes, instead of studying each symptom separately, PsychAD defined 3 NPS contrasts based on those aggregated groups^7^. Therefore, in total, we covered six phenotype contrasts (**Supplementary Note 1.3**): AD vs. Control, AD progression, Pathology-cognition, WeightGain/Sleep/Suicide vs. Control, WeightLoss/PMA vs. Control and Depression/Mood vs. Control.

### Phenotype prediction and cell type prioritization using phenotype association scores of single cells

The phenotype association scores of single cells (PAC scores) calculated by PASCode can be used to predict phenotypes for unseen donors and donors with missing phenotypic labels. This is achieved by taking the averaged PAC scores for each cell type and each donor to construct a donor-by-cell-type feature matrix. Specifically, for *M* donors and *K* cell types, the donor-by-cell-type feature matrix 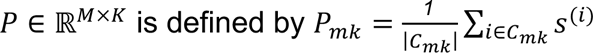, where *s*_*i*_(represents the set of cells from donor *m* that are annotated as cell type *k*, |*C*_*mk*_| denotes the size of the set *C*_*mk*_, and *S*^(*i*)^ is the PAC score for cell *i*. The averaged PAC score reflects the cumulative phenotype association across cells within each cell type for each donor. Subsequently, we fit a Random Forest classifier^16^ (RF) on *P*, which then predicts the phenotype for unseen donors. The trained RF model can also be applied for prioritizing cell types important to particular phenotypes. This is achieved by applying SHapley Additive exPlanations (SHAP)^17^, which takes the RF and the donor-by-cell-type feature matrix *P* as input, and ranks cell types according to their contributions to the RF model accuracy. To ensure statistical robustness and mitigate the effects of random variability in model training, we repeated 100 runs with different random seeds and used their averaged SHAP values.

### Independent validation datasets

Two datasets, SEA-AD^19^ and ROSMAP^20^, were utilized for independent validation. We preprocessed the datasets using Scanpy (v1.9.3). In SEA-AD, to retain donor number balance, we randomly selected 39 donors with dementia and 39 donors without dementia DLPFC region single-cell data.

Furthermore, we transferred the cell type labels from PsychAD (as a reference dataset) to annotate single cells in SEA-AD and ROSMAP (**Supplementary** Fig.5). Refer to **Supplementary Note 2.1** for further details on these preprocessing steps.

### Differential gene expression analysis AD vs. Control

To investigate the importance of inferring PACs over using phenotypes at donor level, two differentially expressed (DE) gene analyses were performed for each cell type. They are donor-level DE gene analysis (i.e., AD vs. Control) and PAC-level DE gene analysis (i.e., *AD*-*PAC*^+^ vs *AD*-*PAC*^−^). It should be noted that our PACs were predicted from both phenotype donors. As an example, for the AD vs. Control contrast, our predictions classify a small number of cells from Control donors as AD- PACs. These cells were removed from the AD-PACs when we performed the PAC-level DE analysis. We only considered the protein-coding genes for the DE gene analyses, and the analyses were conducted separately for each cell type. We used the ‘rank_genes_groups()’ function with the ‘Wilcoxon rank-sum’ method implemented in Scanpy (v1.9.3) to identify DE genes using thresholds of adjusted p-value smaller than 0.05. The obtained DE genes for the AD vs. Control contrast (**Fig. 2, Supplementary** Fig. 2) were evaluated using three methods 1) based on statistical significance, 2) based on AUCell enrichment scores, and 3) based on Gene Set Variation Analysis (GSVA) enrichment. More on these evaluations and the corresponding statistical analysis were detailed in **Supplementary Notes 2.2 and 2.3**, respectively.

#### AD progression

Based on the cell type prioritization depicted in **Fig 3e**, we identified three cell types EN_L2_3_IT, EN_L3_5_IT_2, and EN_L3_5_IT_3 that showed the highest difference across AD-resilient and AD- strict donors. Thus, we conducted DE analysis for these three cell types across their *PAC*^+^cells (i.e., cells that are associated with higher Braak stages). We conducted the DE analysis using MAST^36^ and used Braak stages, sex, and cellular detection rate as covariates. Significant DE genes (i.e., adjusted p-value < 0.05 and |log2(fold change)| >= 0.5) for the three cell types were depicted in **Fig. 3f** and **Supplementary Data 2**.

#### Neuropsychiatric symptoms

We conducted DE analysis for the three neuropsychiatric symptoms, WeightGain/Sleep/Suicide, WeightLoss/PMA (weight loss and psychomotor agitation) and Depression/Mood at cell subclass level using the ‘rank_genes_groups()’ function with the ‘Wilcoxon rank-sum’ method implemented in Scanpy (v1.9.3). Statistically significant DE genes, with adjusted p-value less than 0.05, were provided in **Supplementary Data 7**.

### AD progression stage time

To investigate the gene expression dynamics of cells associated with AD progression, we included a small proportion of donors (n=30) with Braak-stage 0 and donors (n=30) with Braak-stage 6 as the training dataset (**Fig. 3a**, left) to identify AD-progression-PACs (**Fig. 3a**, right) across all Braak stages. A Random Forest classifier was trained on the averaged PAC scores of AD-progression- PACs, which outputs a probability to quantify the donors’ phenotypic similarity with Braak-stage 6 donors based on PACs analysis. The higher the probability is, the more severe the donor’s Braak-stage is. It reflects the pseudotime for AD Braak-stage progression, thus is termed ‘AD progression stage time’.

### Trajectory inference

To further understand the AD progression in terms of Braak stages, we inferred a single-cell trajectory for EN_L2_3_IT using the top 5000 highly variable genes *AD*-*progression*-*PAC*^+^ cells (**Supplementary Data 2**). It should be noted that we isolated only the *AD*-*progression*-*PAC*^+^cells coming from AD-resilient and AD-strict donors with the intention of capturing the contrast effects of pathology-cognition and AD progression together. The highly variable genes were identified using the Seurat (v4.3.0.1) function ‘FindVariableFeatures()’ with the selection method ‘vst’ and 5000 features. Then, these 5000 highly variable genes were scaled and principal components were calculated using the standard Seurat functions. The nearest neighbors were inferred using the top 50 principal components. Then, the UMAP embedding dimensions for the trajectory were estimated and used for trajectory inference using the R package, Monocle3 (v1.3.1)^90^. Using the package Monocle3, we first identified partitions using the ‘cluster_cells()’ function and then inferred the trajectory using ‘lerarn_graph()’ function with modified graph control settings (i.e., euclidean_distance_ratio: 3, geodesic_distance_ratio: 1, minimal_branch_len: 1, orthogonal_proj_tip: False, prune_graph: True, scale: False, rann.k: NULL, maxiter: 20, eps: 10^-5^, L1.gamma: 0.005, L1.sigma: 0.01). Then, the pseudotimes were estimated using Early-AD (Braak 0-2) cells as the root cell group. The root trajectory node, where pseudotime is set to zero, was selected based on the highest percentage of cells coming from the Early-AD cells.

### Differential progression genes in AD resilience

Differential progression of AD-resilient and AD-strict PACs as well as Braak stage groups (**Extended Fig. 3c**) were obtained by binning the cells into 10 groups based on inferred pseudotimes. Using the ‘graph_test()’ function in Monocle3, we identified the trajectory driver genes (i.e., AD progression genes). It used the spatial autocorrelation method, Moran’s I test, to estimate dynamically varying gene expressions with the principal graph. Then, the genes that have q-value < 0.01 and Moran’s I score > 0.1 were isolated (**Supplementary Data 2**) for downstream analysis. Gene set enrichment analysis of these genes was performed to identify related enrichment terms using Metascape^78^ online platform. A custom list of background genes (i.e., top 5000 highly variable genes) was provided for the analysis to ensure the accuracy of enriched terms. The enrichment results are depicted in terms of bubble plots, and all the identified enriched terms are provided in **Supplementary Data 2**. Notably, we overlapped the DE genes identified in **Fig. 3f** with the AD progression genes (i.e., trajectory driver genes) to investigate the AD progression genes differentially expressed across AD-resilient and AD- strict in **Fig. 3h** using a heatmap depicting the averaged pseudo-temporal variation of gene expression.

### Screening for blood inflammation markers

We screened several common blood inflammation marker genes (*SAA1*, *TNF*, *IL1B*, *IL6*, *IL10*, *IL12A*, *IL18*, *IFNG*) by comparing *Depression*-*PAC*^+^with *Depression*-*PAC*^−^ within the same cell subclass.

The Scanpy (v1.9.3) function ‘scanpy.tl.rank_genes_groups’ with Wilcoxon test was applied to identify the p-value and log-fold-change (logFC). Cell subclasses with less than 100 *Depression*-*PAC*^+^ Or *Depression*-*PAC*^−^cells were excluded for comparison.

### Phenotype associated cell subpopulation shared by AD and depression

We used the Scanpy (v1.9.3) function ‘sc.tl.leiden’ with resolution=5 to identify 89 leiden clusters from donors that have both AD and depression information (**Supplementary** Figs. 9**, 10**). Hypergeometric test was performed to determine the significance of overlapping between *AD*-*PAC*^+^and *Depression*-*PAC*^+^within a leiden cluster. Specifically, for each of the 89 clusters, we used python ‘scipy.stats.hypergeom(M, n, N)’, where ‘M’ is the population size that equals the total number of cells within the cluster, ‘n’ represents the number of *AD*-*PAC*^+^ cells within the cluster, and ‘N’ is the number of cells chosen without replacement from the population ‘M’, which equals the number of *Depression*- *PAC*^+^ cells within the cluster. Assuming ‘k’ equals the observed number of cells that are both *AD*- *PAC*^+^and *Depression*-*PAC*^+^, then the p-values were determined as the likelihood of obtaining a greater number of overlapping cells than ’k’ purely by chance which can be modeled by a hypergeometric distribution. Benjamini-Hochberge procedure was used to calculate the FDR values, and only clusters with FDR<0.05 were considered as significant ones shared between *AD*-*PAC*^+^ and

*Depression*-*PAC*^+^.

### Construction of gene regulatory networks

We were interested in applying network biology techniques to better understand dysregulated gene interactions in depression associated astrocytes. To do this, we applied the following procedure to two sets of astrocytes labeled as *Depression*-*PAC*^+^ and *Depression*-*PAC*^−^. First, the log-normalized gene by cell CPM matrix was filtered to retain 5000 highly variable genes identified using the ‘highly_variable_genes()’ function in Scanpy (v1.9.3) library^21^. The gene expression matrices with remaining genes, along with the list of genes annotated as TFs, were supplied to GRNboost2 to infer potentially regulatory links between TFs and target genes based on their expression profiles^91^. From the resulting gene regulatory network, which links TFs to genes, we computed the overlaps between the predicted TFs of every pair of genes. The overlap was quantified as the Jaccard’s Index and arranged as an adjacency matrix (*A*) with values ranging from 0 to 1, essentially depicting gene-gene co-regulation strength. The adjacency matrix was used to find clusters of coregulated genes, or modules, as described below.

### Predicting network modules

The adjacency matrix *A* depicting strength of co-regulation between every pair of genes was utilized to find network modules. First, a dissimilarity matrix was calculated as 1 - *A* and clustered using hierarchical clustering using the average linkage method. The cutreeDynamic function of WGCNA was used to cut the hierarchical clustering dendrogram and find network modules. A minimum module size of 30 was chosen to guarantee enrichment and statistical analysis of the resulting modules. All these operations were performed using the WGCNA package in R^92^. The resulting modules were annotated by statistical enrichments based on disease and gene ontology catalogs, as well as regulatory enrichments based on overlaps with predicted TF targets (**Supplementary Note 3**).

### PASCode web application

We provide an interactive web application for users to explore PACs in this study. Users have custom options for UMAP embeddings, donor phenotypic labels, and PAC scores for different visualization purposes. Specifically, we offer the choice of various UMAP embeddings, including those of gene expression profiles and PASCode latent spaces, for each of the 5 phenotypes. Additionally, users can select donor-level phenotypic labels to color the UMAP, including demographic information such as sex, age, and ethnicity, and AD/NPS phenotypic labels. Five types of PAC scores corresponding to the phenotypes are also available for UMAP coloring. The web application can be accessed at https://daifengwanglab.shinyapps.io/PASCodeDB.

### PASCode pre-trained models

We provide our pre-trained models corresponding to the AD/NPS phenotypes (i.e., AD, AD progression, WeightGain/Sleep/Suicide, WeightLoss/PMA and Depression/Mood) for the community to use. These models were pre-trained on the PsychAD data with million-scale single-cell gene expression profiles, and can be used to annotate the PAC scores for independent single-cell gene expression datasets, as exemplified by our analysis for the AD vs. Control contrast. Tutorials of PASCode and demos on the use of the pre-trained models are provided at https://github.com/daifengwanglab/PASCode.

## Data availability

Raw and processed PsychAD data can be accessed via Sage Bionetworks. Phenotype associated cells and their gene analysis can be interactively visualized at https://daifengwanglab.shinyapps.io/PASCodeDB. Further information on the interactive Web App can be found in **Methods**. All other data is included in the main paper or the Supplementary Data.

All PsychAD data are also available via the AD Knowledge Portal (https://adknowledgeportal.org). The AD Knowledge Portal is a platform for accessing data, analyses, and tools generated by the Accelerating Medicines Partnership (AMP-AD) Target Discovery Program and other National Institute on Aging (NIA)-supported programs to enable open-science practices and accelerate translational learning. The data, analyses and tools are shared early in the research cycle without a publication embargo on secondary use. Data is available for general research use according to the following requirements for data access and data attribution (https://adknowledgeportal.synapse.org/Data%20Access). The results published here are in whole or in part based on data obtained from the AD Knowledge Portal.

## Code availability

The PASCode framework and the pretrained AD/NPS models, together with tutorials and demos can be accessed at https://github.com/daifengwanglab/PASCode. All code and data used for generating figures can be accessed at Zenodo https://zenodo.org/doi/10.5281/zenodo.13241021.

## Acknowledgments

We would like to express our deep gratitude to the patients and their families who generously donated the invaluable biological material essential for the success of this study. We are profoundly indebted to their participation and commitment to advancing scientific knowledge and improving human health. We acknowledge the National Institutes of Health grants, R01AG067025 (to P.R. and D.W.), RF1MH128695 (to D.W.), R21NS127432 (to D.W.), R21NS128761 (to D.W.), U01MH116492 (to D.W.), U01MH116442 (to P.R.), R01MH110921 (to P.R.), R01MH109677 (to P.R.), P50HD105353 (to Waisman Center), National Science Foundation Career Award 2144475 (to D.W.), Simons Foundation Autism Research Initiative pilot grant 971316 (to D.W.), R01MH118827 (to X.Z.), R01MH136152 (to X.Z.), DOD W81XWH-22-1-0621 (to X.Z.), Kellett Mid-Career Award (to X.Z.), Jenni and Kyle Professorship (to X.Z.), and the start-up funding for D.W. from the Office of the Vice Chancellor for Research and Graduate Education at the University of Wisconsin–Madison. The funders had no role in study design, data collection and analysis, decision to publish, or manuscript preparation.

## Author contributions

Conceptualization: DW, PR Methodology: CH, AZL, KHA, DW Software: AZL, KHA, CH

Formal analysis: CH, AZL, KHA, CG, XZ, DW Investigation: XH, XZ, KG, GV, JB, JFF, DL, GEH, PR Resources: DL, JB, KG, GV, GEH, JFF, PR

Data Curation: DL, JB, KG, GV, GEH, JFF

Writing: CH, AZL, KHA, CG, XH, DW, PR with support from all co-authors. Visualization: CH, AZL, KHA, CG

Supervision: DW, PR

Funding acquisition: DW, PR, XZ

All authors read and approved the final draft of the paper.

## Materials & Correspondence

Correspondence to Panos Roussos or Daifeng Wang

## Supplementary Information

### Supplementary materials

Supplementary Notes 1-3, Supplementary Figures 1-12, Supplementary Tables 1-6, Supplementary Data 1-7.

### PsychAD dataset

Description of PsychAD cohort dataset

### PsychAD Consortium Authors

Aram Hong (1, 4, 6, 7); Athan Z. Li (10, 12); Biao Zeng (1, 4, 6, 7); Chenfeng He (9, 12); Chirag Gupta (9, 12); Christian Porras (1, 4, 6, 7); Clara Casey (1, 4, 6, 7); Colleen A. McClung (18); Collin Spencer (1, 4, 6, 7); Daifeng Wang (9, 10, 12); David A. Bennett (19); David Burstein (1, 2, 4, 6, 7, 8); Deepika Mathur (1, 4, 6, 7); Donghoon Lee (1, 4, 6, 7); Fotios Tsetsos (1, 2, 4, 6, 7); Gabriel E. Hoffman (1, 2, 4, 6, 7, 8); Genadi Ryan (13, 17); Georgios Voloudakis (1, 2, 3, 4, 6, 7, 8); Hui Yang (1, 4, 6, 7); Jaroslav Bendl (1, 4, 6, 7); Jerome J. Choi (11, 12); John F. Fullard (1, 4, 6, 7); Kalpana H. Arachchilage (9, 12); Karen Therrien (1, 4, 6, 7); Kiran Girdhar (1, 4, 6, 7); Lars J. Jensen (21); Lisa L. Barnes (19); Logan C. Dumitrescu (22, 23); Lyra Sheu (1, 4, 6, 7); Madeline R. Scott (18); Marcela Alvia (1, 4, 6, 7); Marios Anyfantakis (1, 4, 6, 7); Maxim Signaevsky (6, 7); Mikaela Koutrouli (1, 4, 6, 7, 21); Milos Pjanic (1, 4, 6, 7); Monika Ahirwar (13, 17); Nicolas Y. Masse (1, 4, 6, 7); Noah Cohen Kalafut (10, 12); Panos Roussos (1, 2, 4, 6, 7, 8); Pavan K. Auluck (20); Pavel Katsel (6); Pengfei Dong (1, 4, 6, 7); Pramod B. Chandrashekar (9, 12); Prashant N.M. (1, 4, 6, 7); Rachel Bercovitch (1, 4, 6, 7); Roman Kosoy (1, 4, 6, 7); Sanan Venkatesh (1, 2, 4, 6, 7); Saniya Khullar (9, 12); Sayali A. Alatkar (10, 12); Seon Kinrot (1, 4, 6, 7); Stathis Argyriou (1, 4, 6, 7); Stefano Marenco (20); Steven Finkbeiner (13, 14, 15, 16, 17); Steven P. Kleopoulos (1, 4, 6, 7); Tereza Clarence (1, 4, 6, 7); Timothy J. Hohman (22, 23); Ting Jin (9, 12); Vahram Haroutunian (5, 6, 7, 8); Vivek G. Ramaswamy (13, 17); Xiang Huang (12); Xinyi Wang (1, 4, 6, 7); Zhenyi Wu (1, 4, 6, 7); Zhiping Shao (1, 4, 6, 7)

### PsychAD Consortium Affiliations

1: Center for Disease Neurogenomics, Icahn School of Medicine at Mount Sinai, New York, NY, USA

2: Center for Precision Medicine and Translational Therapeutics, James J. Peters VA Medical Center, Bronx, NY, USA

3: Department of Artificial Intelligence and Human Health, Icahn School of Medicine at Mount Sinai, New York, NY, USA

4: Department of Genetics and Genomic Sciences, Icahn School of Medicine at Mount Sinai, New York, NY, USA

5: Department of Neuroscience, Icahn School of Medicine at Mount Sinai, New York, NY, USA

6: Department of Psychiatry, Icahn School of Medicine at Mount Sinai, New York, NY, USA

7: Friedman Brain Institute, Icahn School of Medicine at Mount Sinai, New York, NY, USA

8: Mental Illness Research, Education and Clinical Center VISN2, James J. Peters VA Medical Center, Bronx, NY, USA

9: Department of Biostatistics and Medical Informatics, University of Wisconsin-Madison, Madison, WI, USA

10: Department of Computer Sciences, University of Wisconsin-Madison, Madison, WI, USA

11: Department of Population Health Sciences, University of Wisconsin-Madison, Madison, WI, USA

12: Waisman Center, University of Wisconsin-Madison, Madison, WI, USA

13: Center for Systems and Therapeutics, Gladstone Institutes, San Francisco, CA, USA

14: Department of Neurology, University of California San Francisco, San Francisco, CA, USA

15: Department of Physiology, University of California San Francisco, San Francisco, CA, USA

16: Neuroscience and Biomedical Sciences Graduate Programs, University of California San Francisco, San Francisco, CA, USA

17: Taube/Koret Center for Neurodegenerative Disease Research, Gladstone Institutes, San Francisco, CA, USA

18: Department of Psychiatry, University of Pittsburgh School of Medicine, Pittsburgh, PA, USA

19: Rush Alzheimer’s Disease Center and Department of Neurological Sciences, Rush University Medical Center, Chicago, IL, USA

20: Human Brain Collection Core, National Institute of Mental Health-Intramural Research Program, Bethesda, MD, USA

21: Novo Nordisk Foundation Center for Protein Research, Faculty of Health and Medical Sciences, University of Copenhagen, Copenhagen, Denmark

22: Vanderbilt Genetics Institute, Vanderbilt University Medical Center, Nashville, TN, USA

23: Vanderbilt Memory & Alzheimer’s Center, Vanderbilt University Medical Center, Nashville, TN, USA

---

**Extended Fig. 1:**
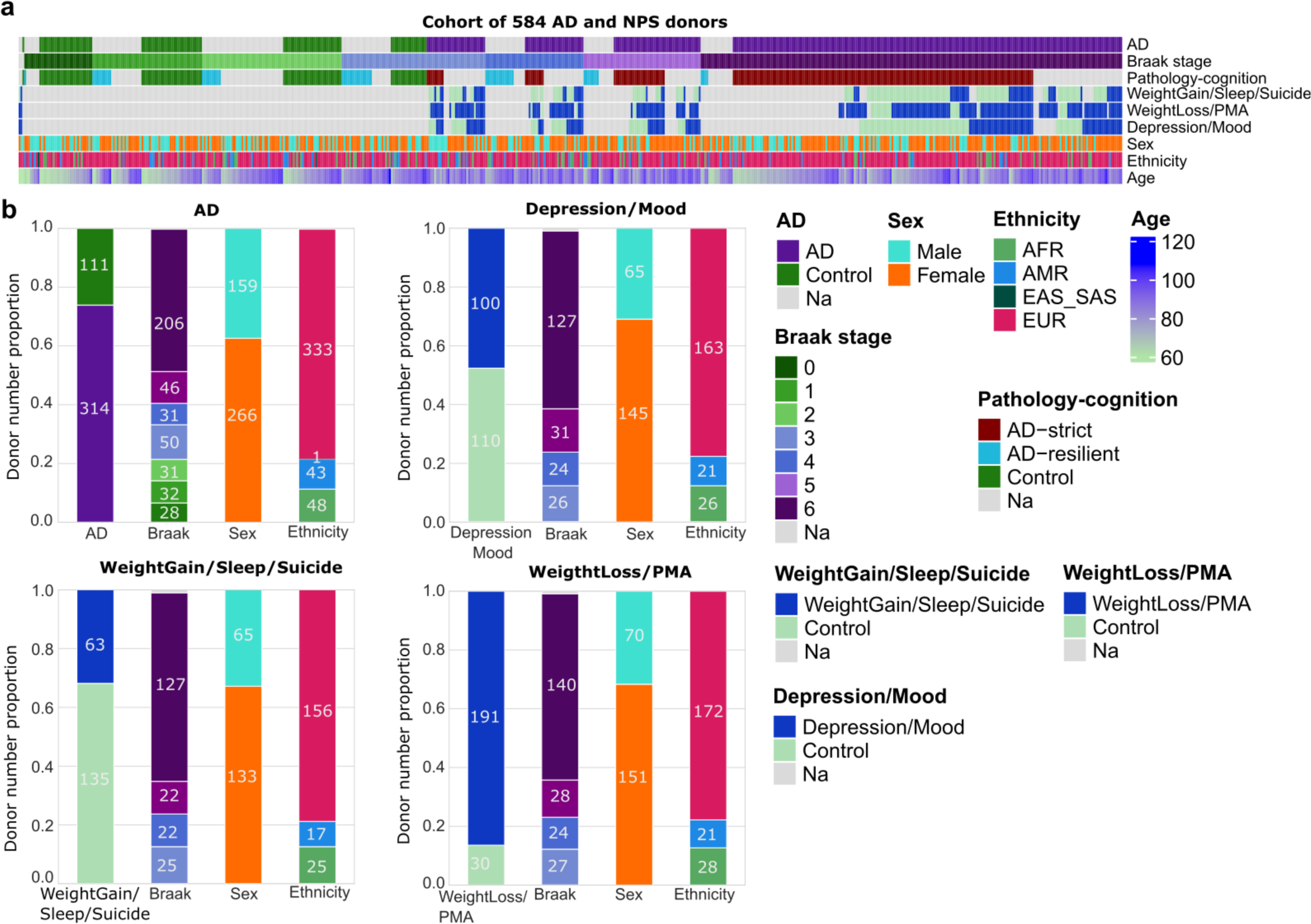
Phenotypic and demographic information of donors from the PsychAD consortium for single-cell phenotypic scoring. a, Columns: Donors (n=584). Rows: Phenotype contrasts and demographic categories (Methods and Supplementary Note 1.3), including AD diagnosis (AD vs. Control), Braak stages (0-6), Pathology-cognition (AD-resilient, AD-strict, and Control), NPS diagnoses (WeightGain/Sleep/Suicide, WeightLoss/PMA forweight loss and psychomotor agitation, Depression/Mood, and Control), Sex (Male vs. Female), Ethnicity (AFR: African; AMR: Admixed American; EUR: European; EAS_SAS: Asian), and Age. Grey color represents unknown labels. **b,** Proportion of donors with AD (upper left), Depression/Mood (upper right), WeightGain/Sleep/Suicide (lower left), and WeightLoss/PMA (weight loss and psychomotor agitation, lower right) across Braak stages, Sex and Ethnicity groups.

**Extended Fig. 2:**
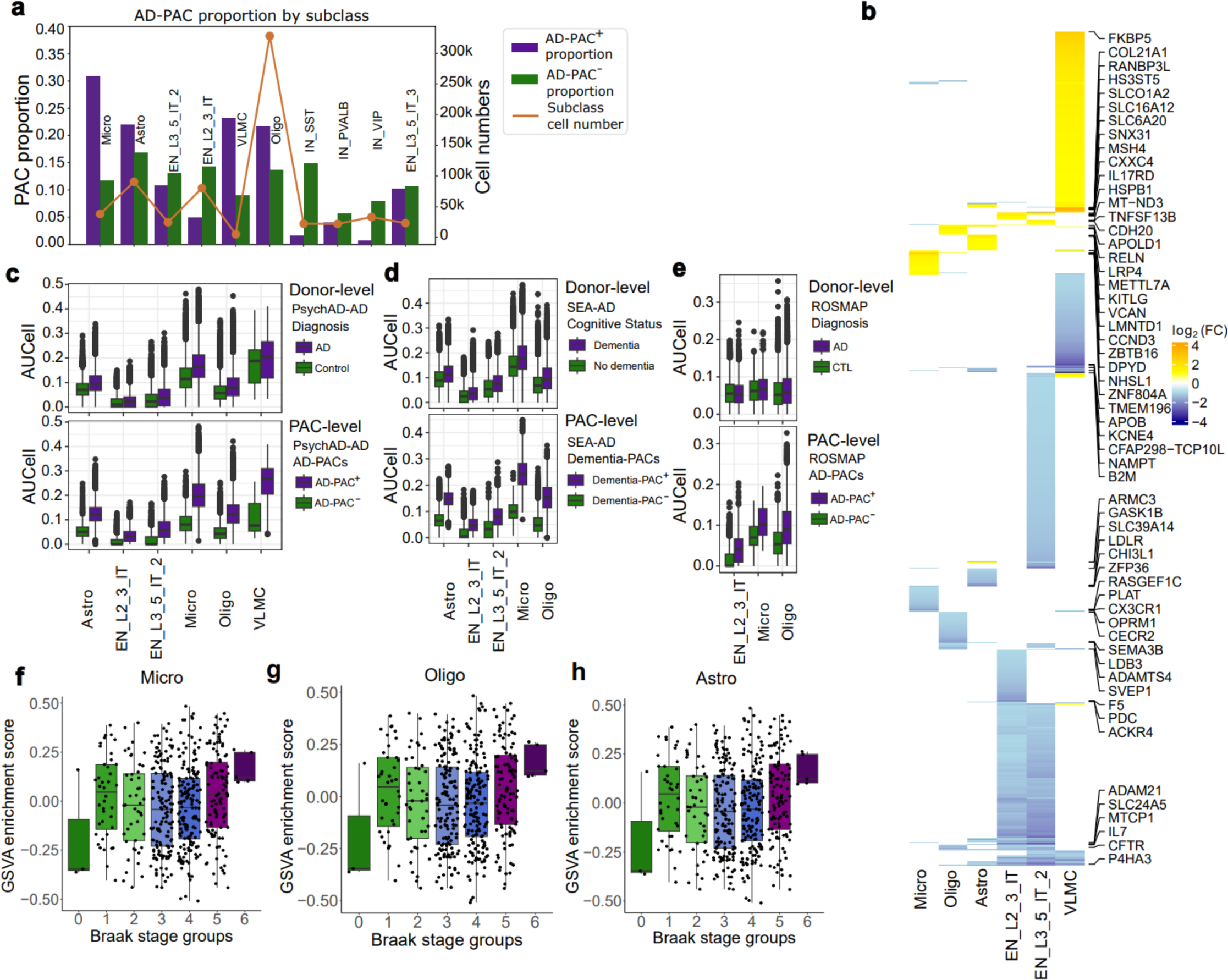
Validation of prioritized Alzheimer’s disease related cell subclasses and genes. a, Left y-axis: Proportion of the numbers of *AD*-*PAC*^+/−^, i.e., cells associated with AD (*AD*-*PAC*^+^) or Control (*AD*-*PAC*^−^) within each cell subclass. Only the top 10 prioritized cell subclasses are displayed (Fig. 2e). Orange curve represents the number of cells within each cell subclass as indicated on the right y-axis. **b,** Differentially expressed (DE) genes between *AD*-*PAC*^+^ and *AD*-*PAC*^−^ within the six top prioritized cell subclasses. **c**, AUCell score distribution for donor-level (top) and PAC-level (below) for PsychAD. AUCell scores were calculated based on the upregulated DE genes of their corresponding cell subclasses in panel **(b)**. Statistical tests to compare (1) AD vs. Control AUCell score distributions at both PAC-level and donor-level (Mann-Whitney U rank test), and (2) the differences observed between AD and Control compared at PAC-level vs. donor-level (bootstrap subsampling), were conducted as described in **Supplementary Note 2.3** and tabulated in **Supplementary Tables 5, 6. d-e,** Similar analysis as in panel **(c)** for independent datasets SEA-AD **(d)** and ROSMAP **(e)**. Several cell subclasses were removed from the SEA-AD and ROSMAP analysis due to the lack of predicted PACs to perform statistical analysis. Related statistics are tabulated in **Supplementary Tables 5, 6**. **f-h,** Gene Set Variation Analysis (GSVA) enrichment of AD bulk RNA-seq dataset^29^ on upregulated DE genes in **(b)** significantly correlated with donor Braak stages for microglia **(f)**, oligodendrocytes **(g)**, and astrocytes **(h)** (Jonckheere-Terpstra trend test p-values <2.42×10^-5^, 8.28×10^-5^, 3.82×10^-6^, respectively). FPKM gene expression values were quantile normalized and batch effect removed as previously described^30^.

**Extended Fig. 3.**
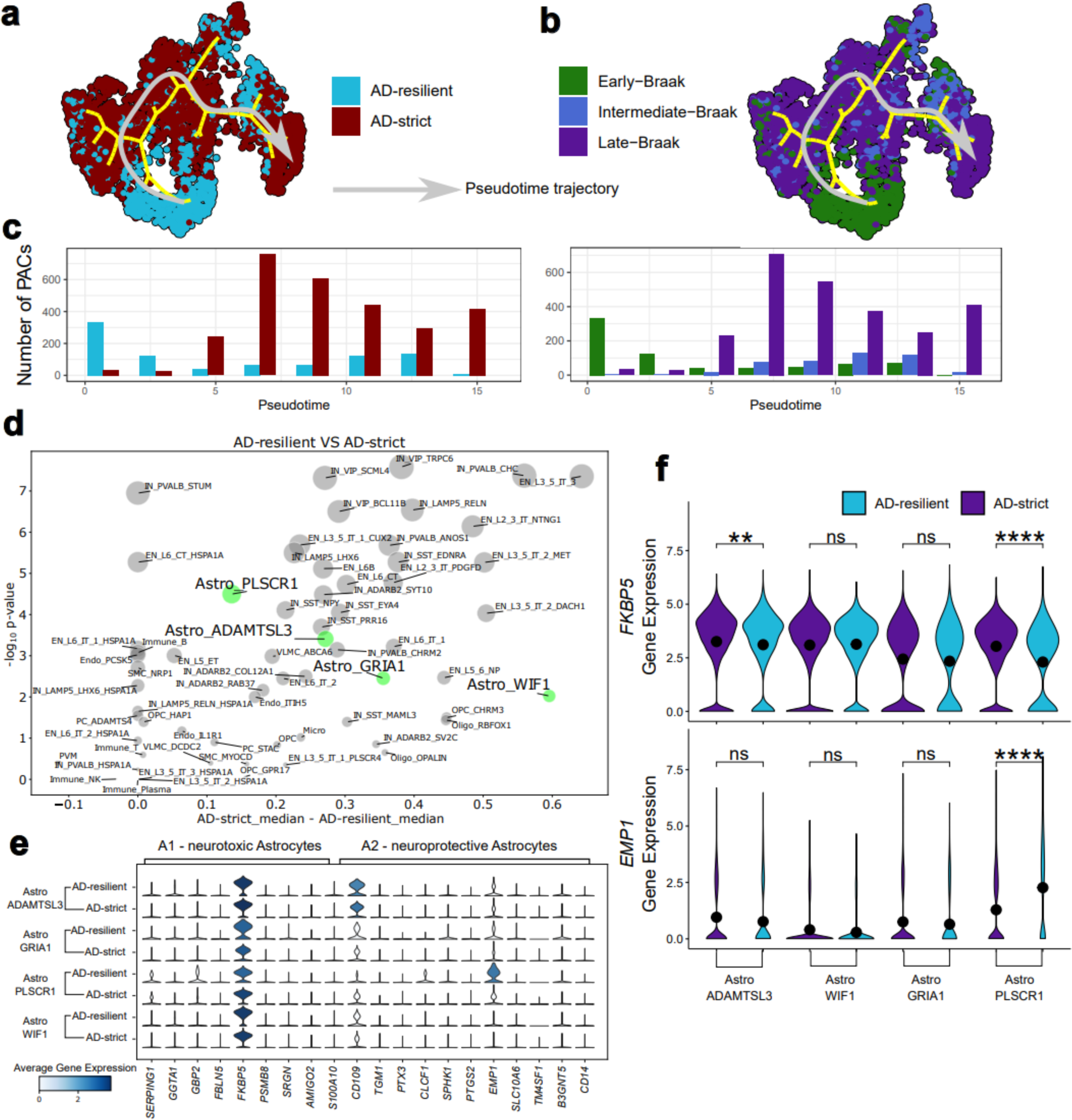
Alzheimer’s disease progression trajectory analysis using AD-progression-PACs within EN_L2_3_IT cells and the role of reactive Astrocytes in cognition. a-b, EN_L2_3_IT cell trajectory colored by AD-resilient/strict (a) and Braak stage (b) (Early-Braak: 0-2, Intermediate-Braak: 3-4, Late-Braak: 5- 6). c, Bar plots showing the differential progression of AD-resilient/strict (left) and Braak stage (right) along pseudotime (Methods). d, Prioritizing cell subtypes that are most different across AD-resilient and AD-strict donors. Only donors with Braak stages 4, 5 and 6 were considered. Both Y-axis and the bubble size represent minus log10 Wilcoxon rank-sum test p-values comparing the SHAP values. To exclude stochastic effects, we trained 100 Random Forest models with different random seeds, and averaged their SHAP values for comparison. X-axis is the difference in the median values of averaged AD-progression-PAC scores between AD-resilient and AD-strict donors. Astrocyte subtypes are colored in green. e, Gene expression markers of neurotoxic (A1) and neuroprotective (A2) reactive astrocytes across astrocyte subtypes and pathology- cognition. f, Violin plots for *FKBP1* and *EMP1* genes and their statistical significance across AD-resilient and AD-strict donors. The pairwise comparisons were conducted with the Wilcoxon rank-sum test.

**Extended Fig. 4.**
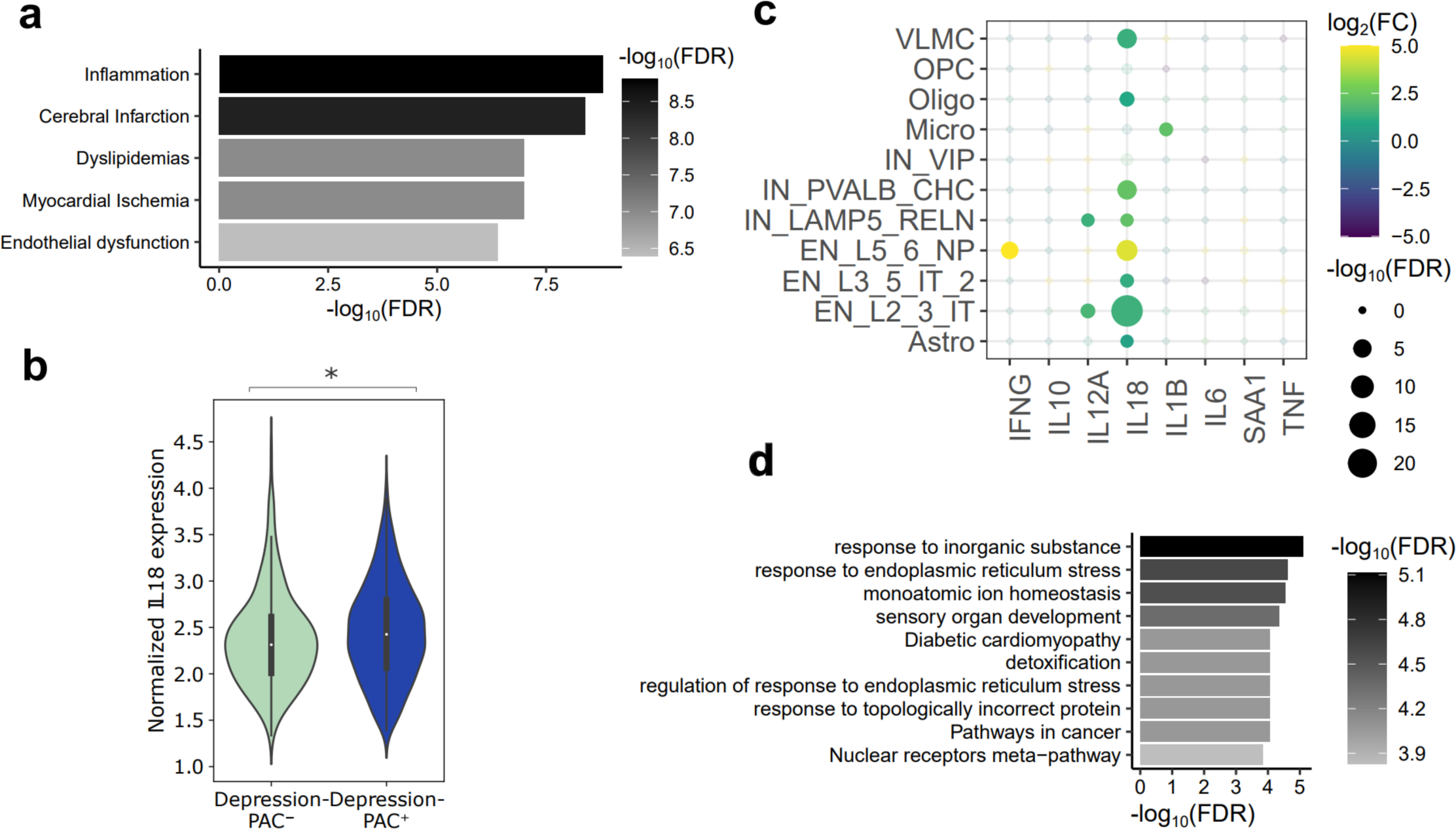
Gene and GO enrichment analysis of depression associated astrocytes in AD. **a**, GO enrichment analysis of significantly upregulated genes (log2 fold change > 0.5 and adjusted p-value < 0.05) within *Depression*-*PAC*^+^(i.e., cells associated with depression) compared with *Depression*-*PAC*^−^(i.e., cells associated with control) within astrocytes. **b,** Bar plot depicting the *IL18* gene expression difference across *Depression*-*PAC*^+^ and *Depression*-*PAC*^−^. Significant difference observed within astrocytes (p < 3.24×10^-3^ based on one-sided Wilcoxon rank-sum test). **c**, Common inflammation markers differentially expressed between *Depression*-*PAC*^+^ and *Depression*-*PAC*^−^ cell subclasses, p-values and fold changes are shown. Cell subclasses with over 100 *Depression*-*PAC*^+^ and *Depression*-*PAC*^−^ are shown. **d**, GO enrichment analysis (**Supplementary Data 3**) of genes significantly upregulated in ‘AD-Depression-AstroWIF1-cluster’ of Fig. 4g compared with all the other Astro_WIF1 cells. GO enrichment was determined with Metascape ^78^.

