## Supplementary Materials for "Phenotype Scoring of Population Scale Single-Cell Data Dissects Alzheimer’s Disease Complexity"

### Supplementary Notes

### Supplementary Note 1: Supplementary Note for "Phenotype scoring of single cells"

We developed Phenotype Associated Single Cell encoder (PASCode), a deep learning framework for scoring phenotype association of cells. PASCode employs an ensemble approach to integrate multiple differential abundance (DA) methods and uses a graph attention network (GAT) for single-cell phenotype association score (PAC score) predictions. In this section, we provide an introduction to the PASCode framework.

#### Supplementary Note 1.1: PASCode for single-cell phenotype scoring

PASCode assigns each cell a PAC score ranging from -1 to 1 to quantify its association with a specific phenotype. Given one phenotype contrast between a positive condition and a negative condition, a score greater than zero indicates an association with the positive condition, while a score less than zero indicates an association with the negative condition. The absolute value of the score indicates the strength of the association, with 1 being the most significant, and 0 being the most insignificant. Such associations can be considered as enrichment of cell subpopulations in regions within the transcriptomic space, with a higher positive PAC score suggesting the cell is located in a region more enriched with the specified positive phenotype condition and vice versa[1]. PAC scores are predicted via a GAT model, which can be trained using the gene expression value to predict the ensembled ‘aggregated phenotype labels’ of several independent differential abundance (DA) methods.

##### Supplementary Note 1.1.1: Select and execute differential abundance methods

**Supplementary Table 1** summarizes several state-of-the-art DA methods, we explain whether and why certain methods have been selected to be integrated into PASCode. As in the table, several DA methods based on clustering[2–4] involve grouping transcriptomics-ally similar cells into clusters to identify those with cell proportions significantly altered with phenotype condition changes. However, these clustering-based methods are suboptimal, especially when the altered cells do not fall into well predefined subclusters, such as those cell stages during  continuous differentiation[5,6]. Several other DA methods are clustering-free, which aim to identify DA cells by directly zooming into cell-level without relying on predefined clusters[5,7–10]. In this work, clustering-free DA methods were chosen over clustering-based ones due to their cellular-level resolution and superior performance for disease gene identification. The selected clustering-free methods (hereafter referred to as “DA methods”) all rely on the cell-cell similarity graph for DA cells identification. Specifically, k-nearest neighbors (KNN) and Uniform Manifold Approximation and Projection[11] (UMAP) are the main graph construction algorithms used, which are performed within the principal component spaces (PC=50). Each method takes the cell-cell similarity graph and donor-level phenotype labels as input, producing a specific measurement of abundance as output, which will then be input to an aggregation algorithm for obtaining single-cell ‘aggregated phenotype labels’ for GAT training (**Supplementary Note 1.1.2**). Details about parameters choices of the selected DA methods are provided in **Supplementary Table 2**.

#### Supplementary Note 1.1.2: Rank aggregation for robust single-cell aggregated phenotype label identification

An ensemble approach that integrates the outputs of multiple DA methods would potentially improve the accuracy. To integrate those selected DA methods in an unbiased manner, we employed the Robust Rank Aggregation[12] (RRA) algorithm. Each DA method can generate a list of cells ranked by the method's specific measurement. RRA finds cells that are preferentially highly ranked across multiple lists, a p-value-based score is assigned by comparing across the ranks against a null hypothesis assuming those ranks follow a uniform distribution. Notably, the aggregation is purely rank based, so is robust to the distribution of individual method's score distribution (**Supplementary Fig. 12**). We aggregated the positive and negative associations separately by cutting each method's scores at zero. Within the positive (negative) portion, RRA identified cells consistently ranked high (low) across multiple lists by comparing them with a uniform distribution. Afterwards, a p-value can be calculated, with a small p-value indicating low probability of consistent ranking by chance. After the positive and negative ranks were aggregated separately, p-values from the negative portion were reversed (i.e. times by -1) and joined with p-values from the positive one. Notably, the RRA aggregation generated a sparse distribution of the p-values, which resulted in a deterministic identification of cells positively/negatively associated with certain phenotypes **(Supplementary Fig. 12)**. Without losing generality, we chose +0.5 and -0.5 as thresholds to assign cells into three 'aggregated phenotype labels' from RRA aggregation results: 'positive aggregated label' represents cells significantly associated with the positive condition of the phenotype; cells with 'negative aggregated label' are significantly associated with the negative condition; cells with 'non-labels' have an insignificant phenotype association with neither of the conditions.

We benchmarked the accuracy of several DA methods along with RRA with synthesized datasets (**Supplementary Note 1.2.1** and **Supplementary Fig. 1-2**), and found that RRA outperforms any single method alone **(Supplementary Fig. 1)**. We then selected methods with the highest accuracy (i.e., Milo[7], MELD[8] and DAseq[5]) for aggregation to ensure the most robust RRA integration throughout this work.

#### Supplementary Note 1.1.3: GAT for phenotype scoring of single-cells

Despite the fact that RRA achieves a higher performance for identifying phenotype associated cells than any individual DA method, it suffers from several challenges when applied to population-level datasets. Firstly, similar to DA methods, it is prone to data imbalance in terms of donor phenotypes (**Supplementary Fig. 2**), which are common in population-level datasets. Secondly, most DA methods (e.g., MELD, DAseq) are computationally expensive, leading to prolonged running times for large datasets. Thirdly, since DA methods identify cells whose abundances significantly change in response to donor phenotypes, they all require the phenotypes as input for quantifying single-cell phenotype association, so they can not handle donors with missing phenotypes.

To address these challenges, PASCode follows the approach for training a machine learning model, which is to learn the gene expression patterns for different 'aggregated phenotype labels' within one dataset, and to predict labels for unseen cells in others. Since the machine learning models were pre-trained on balanced datasets, it would potentially mitigate the reduction of performance for phenotype associated cells identification on imbalanced phenotypes (**Supplementary Note 1.2.1, Supplementary Fig. 2**). Additionally, the pre-trained models allow predicting phenotype labels for cells from unseen donors, even in the presence of missing phenotypes or when using independent cohorts, as demonstrated by the ROSMAP and SEA-AD atlas in **Fig. 2** and **Fig. 3**.

Similarities among cells can be modeled using cell-cell graphs. Graph representation captures the heterogeneities of single-cell data and accounts for the similarity information among cells (e.g. cell type differences), and is commonly used for single-cell analyses[13]. Hence, we employed Graph Attention Network (GAT)[14] that takes cell-cell KNN graphs and gene expression data as input to predict the RRA aggregated phenotype labels. We argue that GAT makes use of cell-cell similarity information in the transcriptomic space, thus would achieve high accuracy by paying attention to the local cell-cell similarity. Benchmarking experiments also showed that GAT outperformed other common machine learning models, including random forest[15], multilayer perceptron, and graph convolutional networks (**Supplementary Fig. 3**). We implemented the GAT model using the PyTorch[16] and PyG[17] framework.

**Graph construction.** We employed the python package Scanpy (v1.9.3)[18] for cell-cell KNN graph construction, which is widely applied in the single-cell analysis community. First, the algorithm computed a number of principal components (PCs) from gene expression data. Then, a cell-cell connectivities matrix was calculated using UMAP in the PC space. In particular, we utilized 'scanpy.pp.pca()' followed by 'scanpy.pp.neighbors()' with default parameters (number of neighbors= $15$, number of PCs=$50$) to generate a cell-cell connectivities matrix. The matrix was then converted into an adjacency matrix (with binary entries indicating whether two cells were connected in the graph) as input to the GAT model.

**Graph Attention Network.** The inputs of GAT are a set of $N$ nodes, each representing a cell, where each node/cell $i$ ( $i = 1, ..., N$) has a gene expression feature vector $x^{(i)} \in \mathbb{R}^d$ and a set of $E$ undirected edges, with each edge connecting two nodes. The neural network processes the feature vector $x^{(i)}$ of each node through several graph attention layers. The output from the neural network is passed through a softmax activation function, resulting in a probability distribution over three values $\hat{y}^{(i)} \in \mathbb{R}^3$ . Here, the $j$th value $\hat{y}_j^{(i)}$ represents the predicted probability of node/cell $i$ being assigned to class $j \in \{1, 2, 3\}$: $PAC^-$ as class 1, Non-PAC as class 2, and $PAC^+$ as class 3. We use cross entropy loss as the objective function for model training:

$$\mathcal{L} = -\sum_{i=1}^{N}\sum_{j=1}^{3} y_j^{(i)} log(\hat{y}_j^{(i)}) ,$$

where $y^{(i)}$ is the ground truth one-hot encoded vector representing the class label of node/cell $i$: $PAC^-$ $[1, 0, 0]$; Non-PAC $[0, 1, 0]$; $PAC^+$ $[0, 0, 1]$.

For simplicity in ranking aggregation, we utilized a single PAC score $s^{(i)}$ as a clear and comparable metric to represent a cell's phenotype association, where $s^{(i)} = \hat{y}_3^{(i)} - \hat{y}_1^{(i)}$ and $-1 \leq s^{(i)} \leq 1$. This subtraction emphasizes the difference in probability between the cell being associated with positive ($PAC^+$) or negative condition ($PAC^-$), directly correlating the score with the cell's phenotype association strength.

The GAT model comprises three layers: an input layer, a hidden layer with 64 neurons, and an output layer with three neurons. It incorporates a multi-head attention mechanism with four heads to capture the heterogeneities and complexities of single-cell data. We used 90% donors for training and

10% donors for validation, with equal numbers of donors with positive and negative phenotype conditions and balanced male/female ratio in both sets. Training involved an Adam optimizer with 0.001 learning rate and 0.001 weight decay, adjusted by a plateau scheduler with a decay factor of 0.5 every 2 epochs. An early stopping mechanism with patience=10 was implemented to prevent overfitting. All computations were performed on a machine with 256GB RAM and a 24GB RTX A6000 GPU.

##### Supplementary Note 1.1.4: Group cells into PACs based on PAC scores

For the majority of the PAC based analysis in this work, we used the continuous GAT output score (PAC score). However, for analyses that require discrete categorization of cells, such as differential gene expression analysis, we applied +/- 0.5 as the cutoffs. Cells with a PAC score greater than 0.5 were classified as $PAC^{+}$, those with a score less than -0.5 as $PAC^{-}$, and cells with scores ranging from -0.5 to 0.5 were categorized as Non-PAC.

#### Supplementary Note 1.2: Benchmark DA methods and machine learning algorithms

##### Supplementary Note 1.2.1: Benchmark DA methods

Inspired by several recent studies[7,19], we first evaluated selected DA methods with synthetic datasets. Specifically, using the single-cell transcriptomic data from ROSMAP single-cell atlas[20], we simulated two datasets and conducted benchmark experiments in similar ways as detailed in a recent study[1].

The first dataset (sim-data1) was generated in a similar manner as described in Dann et al.[7] to mimic the differential abundance of cells across two conditions (cond1 and cond2). Briefly, raw gene expression data were downloaded and preprocessed (i.e., normalization, scaling, etc.). We then selected the top 2000 highly variable genes and performed leiden clustering using the function 'scanpy.tl.leiden()' (with resolution=0.5) to get 18 clusters as shown in **Supplementary Fig. 4**. We then randomly picked two clusters as ground truth, of which one cluster was assigned as positively associated with cond1 (clusters_pos) while the other as negatively associated with cond1 (i.e. positively associated with cond2, clusters_neg). All the other clusters were assigned as associated with neither cond1 nor cond2 (clusters_non). We then used a probability $p$ ($p > 0.5$) to control the simulated positive/negative/non association of each cluster with cond1. Specifically, cells within clusters_pos were assigned as *cond1* label with probability $p$, cells within clusters_neg were assigned as *cond2* with probability $p$. Cells within clusters_non were randomly assigned as *cond1* or *cond2* with equal probability. To simulate the extent of abundance difference (i.e. the fold change of cond1 vs cond2 cells within clusters_pos and clusters_neg compared with cluster_non), we varied $p$ to 0.7, 0.8 and 0.9. To exclude the effects of specific clustering structures, we repeated this whole process $5$ times by selecting different pairs of clusters as ground truth. Together, each of the five cluster pairs (1,2), (1,3), (4,14), (6,9), (0,11) in **Supplementary Fig. 4** was picked as the ground truth. To exclude the effects of donor variances, cells were also randomly assigned to 48 donors, among which 24 donors were *cond1* and the other 24 were *cond2*. The benchmarking result was shown in **Supplementary Fig. 1**, which demonstrated that RRA outperforms other methods in terms of both accuracy and robustness, as RRA showed more consistent high performances across different $p$ and ground truth cluster choices.

The second dataset (sim-data2) was simulated to test the influence of donor imbalance on the selected DA methods, RRA, and GAT. Unlike sim-data1, $p$ was fixed as 0.9 and one *cond1*/*cond2* pair was pre-selected. To simulate the imbalance of donors, we gradually remove donors with *cond1* or *cond2* labels. Together, the following donor ratios between *cond1* vs *cond2* donors: 2vs24, 6vs24, 12vs24, 18vs24, 24vs18, 24vs12, 24vs6, 24vs2 were used to account for various donor imbalance. Experiment results were summarized in **Supplementary Fig. 2**, which shows that single DA methods were susceptible to imbalanced donor numbers across conditions. The result also showed RRA mitigated this issue by outperforming other single DA methods in the donor imbalance settings. This justified the necessity for subsampling donors and using the donor-number-balanced aggregated labels to train a machine learning model, and then to predict PAC scores of the remaining ones.

To evaluate the performance of each method by comparing with the ground truth labels, we assigned each cell with categorical prediction labels (positive-DA, negative-DA or Non-DA) from the outputs of each method similar to a recent study[1]. For Milo, we chose a spatial-FDR threshold 0.1 to distinguish DAs (either positive or negative DAs) versus Non-DAs, afterwards we used log fold change to separate positive-DAs with negative-DAs, as described in the original work. For MELD, we used the gaussian mixture model according to their tutorial for identifying DAs as suggested by the authors. For CNA, we chose a FDR threshold 0.1 to distinguish DAs versus Non-DAs, and the neighborhood coefficient was used to separate positive-DAs with negative-DAs, according to the tutorial. For DAseq, we directly used their own output, of which the thresholds were determined by random permutation test within the tool.

##### Supplementary Note 1.2.2: Benchmark machine learning algorithms

To find an optimal machine learning algorithm for RRA aggregated phenotype labels prediction, we benchmarked graph attention network (GAT), graph convolutional network, multi-layer perceptron, and random forest[16] using ROSMAP dataset[20] with 6-fold cross-validation. For one fold, the whole dataset of 48 donors was splitted into training and validation sets by a 5:1 ratio, and in each set the AD and sex phenotypes were evenly distributed. We used the aggregated phenotype labels obtained by running the DA methods and RRA on the whole dataset as the “ground truth” labels for evaluations. Common metrics for machine learning classifiers, such as F1-score, precision etc., were calculated for model accuracy evaluation. The result demonstrated the superiority of GAT for aggregated phenotype label prediction (**Supplementary Fig. 3**), as GAT consistently achieved the best scores across various metrics.

#### Supplementary Note 1.3: Glossary of Terms

This section provides the definition and detailed information of covered phenotype contrasts in this work. CERAD scores, semi-quantitative measurements of AD progression, were used for AD diagnosis of the donors and have the values of 1, 2, 3 and 4, representing No AD, Possible AD, Probable AD, and Definite AD, respectively. We considered the following 3 disease-related phenotype contrasts in this work:

##### AD vs. Control

This contrast compares donors diagnosed with Alzheimer’s disease and their controls. Donors with AD were defined as those who have CERAD scores of 2, 3, or 4, Braak stage of 3 and above, and clinically diagnosed dementia. Their respective control group was defined as CERAD score of 1 and Braak stage within 0-3.

##### AD progression

This contrast compares AD progression via Braak stages that measure cortical spread of neurofibrillary tangles, irrespective of donors clinical diagnosis.

##### Pathology-cognition

This contrast integrates the pathological (tau proteinopathy) and cognitive (severity of cognitive decline) information together to group donors into three categories: (1) Donors with a CERAD score of 4, Braak stages above 3, and clinically diagnosed dementia (AD-strict), (2) Donors could have CERAD scores of 2, 3, or 4 and must not have dementia (potential AD resilience or AD-resilient), and (3) Donors with a CERAD score of 1 and Braak stage within 0-3 (Control).

##### WeightGain/Sleep/Suicide vs. Control

These neuropsychiatric symptoms (NPS) correspond to sleep issues (early-, mid-, and late- insomnia, and hypersomnia), weight gain, suicidal ideations, delusional worthlessness, worthlessness, and psychomotor retardation. One donor was considered to be ‘Case’ - if at least one of the above symptoms appeared to be true and ‘Control’ if none of the symptoms was true.

##### WeightLoss/PMA vs. Control

This contrast covers the following NPSs diagnosed in AD donors: weight loss, decreased appetite, psychomotor agitation, loss of energy and ruminations. One donor was considered to be ‘Case’ - if at least one of the above symptoms appeared to be true and ‘Control’ if none of the symptoms was true.

##### Depression/Mood vs. Control

This contrast corresponds to depression and other symptoms associated with mood (dysphoria, anhedonia). One donor was considered to be ‘Case’ - if at least one of the above symptoms appeared to be true and ‘Control’ if none of the symptoms was true.

### Supplementary Note 2: Supplementary Note for "Prioritizing cell subpopulations and genes in AD"

#### Supplementary Note 2.1: Preprocessing of the independent validation datasets

We applied standard Scanpy (v1.9.3) functions for the preprocessing of SEA-AD[21] and ROSMAP[20] data. For SEA-AD, we selected the DLPFC regions of 39 donors with dementia and 39 donors without dementia to retain donor number balance. Cell-cell graph was input to the DA tools to get cell aggregated phenotype labels, which was constructed by UMAP based on the scVI embeddings (n=20) obtained from the original SEA-AD data (‘scanpy.pp.neighbors()’) with default parameters. For GAT predictions, we included the 3,322 genes intersected between SEA-AD (36,601 genes) and PsychAD (3,401 genes), with the rest 79 (3401-3322=79) genes filled with 0.

For ROSMAP single-cell RNA-seq data preprocessing, we first removed genes expressed in less than 3 cells, reducing the number of genes from 17,926 to 17,775. We then performed log-normalization on the expression data with a target sum of 10,000 (scanpy.pp.normalize_total()

with target_sum=1e4, and scanpy.pp.log1p() with default parameters), selected 5,000 highly variable genes (scanpy.pp.highly_variable_genes() with n_top_genes=5000), and performed standard scaling (scanpy.pp.scale() with default parameters). The cell-cell graph was constructed using UMAP with 50 PCs of the highly-variable-gene expression data, and was then input to the selected DA tools (Milo, MELD, and DAseq) to get aggregated cell labels. Similar to SEA-AD, for GAT predictions, we included the 3,218 genes intersected between ROSMAP (17,926 genes) and PsychAD (3,401 genes), with the rest 183 (3401-3218=183) genes filled with 0.

For donor level phenotype prediction and cell type prioritization, we applied the function ‘scanpy.tl.ingest()’ for label-transferring to ensure the SEA-AD and ROSMAP cells to have the same cell type annotation as PsychAD (**Supplementary Fig. 5**).

### Supplementary Note 2.2: Differentially expressed genes comparison at donor and PAC levels

#### Comparison by statistical significance

A scatter plot indicating the statistical significance of DE gene analyses at donor and PAC levels for Microglia is shown in **Fig. 2g**. First, the genes with absolute $\log_2$(fold change) greater than 0.5 were identified in both DE analyses and overlapped to find conserved and specific DE genes. To show the relationship of these identified DE genes with AD, we cross-checked them with genes from three known Microglia AD-related enrichment terms (i.e., GO:001540, Amyloid-beta binding; GO:0048156, Tau protein binding; KEGG: hsa05010, Alzheimer disease-Homo sapiens) in a public database from Davis et al.[22]. To ensure the statistical significance of DE genes at PAC level is always greater than at the donor level, we extended our analysis to Oligodendrocytes, Astrocytes, EN_L2_3_IT, and EN_L3_5_IT_2 cell subclasses. We used a $\log_2$(fold change) greater than 1.5 as a cutoff for identification of DE genes (**Supplementary Fig. 7a-d**).

#### Comparison by AUCell scores

We used AUCell (v1.12.2) scores[23] to investigate the identified DE genes and AD genes (i.e., GO:001540, Amyloid-beta binding; GO:0048156, Tau protein binding; KEGG: hsa05010, Alzheimer disease-Homo sapiens). Since the genes curated from the three AD-related enrichment terms consist of both up- and down-regulated genes, we overlapped them with the upregulated genes in Microglia reported by Mathys et al.[20]. Then, boxplots were used to visualize the distribution of AUCell scores calculated for Microglia cells at both donor and PAC levels **(Fig. 2f)**.

To ensure the robustness of our identified DE genes in our training dataset are similarly enriched in the entire dataset and within other independent AD scRNA-seq datasets (i.e., ROSMAP, SEA-AD), we compared their AUCell scores distribution in Microglia, Astrocytes, and Oligodendrocytes **(Extended Fig. 2c-e).** Notably, we only used the upregulated genes (adjusted p-value < 0.05 and $\log_2$(fold change) > 1, see **Extended Fig. 2b**) to calculate the AUCell scores.

#### Comparison by GSVA enrichment

The FPKM values of the bulk RNA-seq dataset were downloaded as reported by De Jager et al.[24]. Quantile normalization and batch error correction were performed on the downloaded dataset. Gene Set Variation Analysis (GSVA) (v1.50.0)[25] was then applied to calculate the gene expression enrichment on the pre-selected gene-sets. For analysis in **Extended Fig. 2f-h, Supplementary**

**Figure 7e-g**, gene-sets were defined as the upregulated genes within $PAC^{+}$ compared with $PAC^{-}$ for each cell type respectively.

#### Supplementary Note 2.3: AUCell scores based statistical tests

We conducted two statistical tests: (1) to check the estimated AUCell scores are statistically enriched in the $AD\text{-}PAC^{+}$ compared to the $AD\text{-}PAC^{-}$; (2) to check whether the AD vs. control difference observed at the PAC level is statistically higher compared to the difference observed at the donor level. We first used the estimated AUCell scores for the three AD-related enrichment terms (**Fig. 2f, Supplementary Note 2.2, and Supplementary Table 3**) in Microglial cells to compare the AD and control cells using Mann-Whitney U rank test. We used the 'mannwhitneyu()' function in python Scipy (v1.11.1) package for the analysis. The p-values and their observed mean AUCell value differences across the two groups (i.e., AD and Control) are provided in **Supplementary Table 3**.

Further comparisons were made to investigate the performance of PACs in terms of capturing AD related genes. Here, we conducted a bootstrap resampling 1000 times with 75% of randomly sampled cells to calculate the mean difference across AD and control at both the PAC and donor levels. Then the mean differences were compared using the Mann-Whitney U rank test. **Supplementary Table 4** provides the corresponding p-values and the mean differences observed at PAC and donor levels.

Similarly, we conducted the two sets of statistical tests for the AUCell enrichment study of PAC based upregulated genes estimated for Astrocytes, Oligodendrocytes, EN_L2_3_IT, EN_L3_5_IT_2, VLMC and Microglia in three independent validation datasets (**Supplementary Note 2.2, Extended Fig. 2c-e,** and **Supplementary Data 1**). The statistics of pairwise comparisons and PAC vs Donor-level comparisons were included in **Supplementary Table 5** and **6**, respectively.

### Supplementary Note 3: Supplementary Note for "Astrocyte gene networks in depression"

#### Module annotation

Note that our approach of linking target genes to each other and predicting modules is different from a standard WGCNA type approach. In our approach, we measure the expression relationships of TFs and their potential targets. This approach lets us harness the regulatory potential of TFs that do not have their binding sites known yet. Furthermore, it is also important to note that our module analysis is different from modules defined by a popular GRN inference tool called SCENIC. The main difference between the two techniques is that our strategy is able to link target genes to each other, while SCENIC reports regulons (TF→gene relationships) in which non-TF genes are not explicitly connected to each other, and the output remains devoid of common TFs for a given target gene pair. Such a structure of GRNs predicted by SCENIC is essentially a bipartite graph, not readily accessible to traditional graph clustering algorithms. Therefore, we chose to use SCENIC as an GRN additional layer to our coregulation modules. For this, we processed the GRNboost2 outputs through the SCENIC pipeline to remove TFs without binding motifs and indirect targets. In the resulting GRNs, we tested the statistical enrichment (as described below) of every regulon (targets of a TF) within each coregulated gene module. TFs that were found to have an over-representation of regulons within coregulated modules were declared as the regulators of those modules.

To gain a better understanding on the functional properties of the predicted gene modules, we tested for enrichment of biological process terms in the human Gene Ontology (GO), disease ontology (DO), and other pathway databases. We removed terms that annotate more than 500 or less than 10 genes as generic and non-informative terms, respectively. The enrichment of remaining terms within each module were statistically evaluated using the hypergeometric tests using the hypeR package in R. The p values from these tests were corrected for multiple testing and enrichments with FDR < 0.1 are reported. Geneset files used in enrichment analysis were obtained from the Enrichment map database[26].

To add TFs as potential regulators of the predicted modules, we tested the enrichment of SCENIC filtered regulons within the modules using a similar statistical test as above. TFs with targets significantly enriched within the modules were labeled as the regulators of the modules.

### TF rewiring analysis

We utilized the GRNboost outputs linking TFs to target genes to analyze rewiring of TFs between $Depression\text{-}PAC^{+}$ versus $Depression\text{-}PAC^{-}$ cells. For each TF, we calculated the difference in edge importance scores between $Depression\text{-}PAC^{+}$ and $Depression\text{-}PAC^{-}$ networks. We then sorted TFs based on the descending order of the absolute difference in edge importance scores and selected the top decile TFs as the most rewired.

### Supplementary Figures

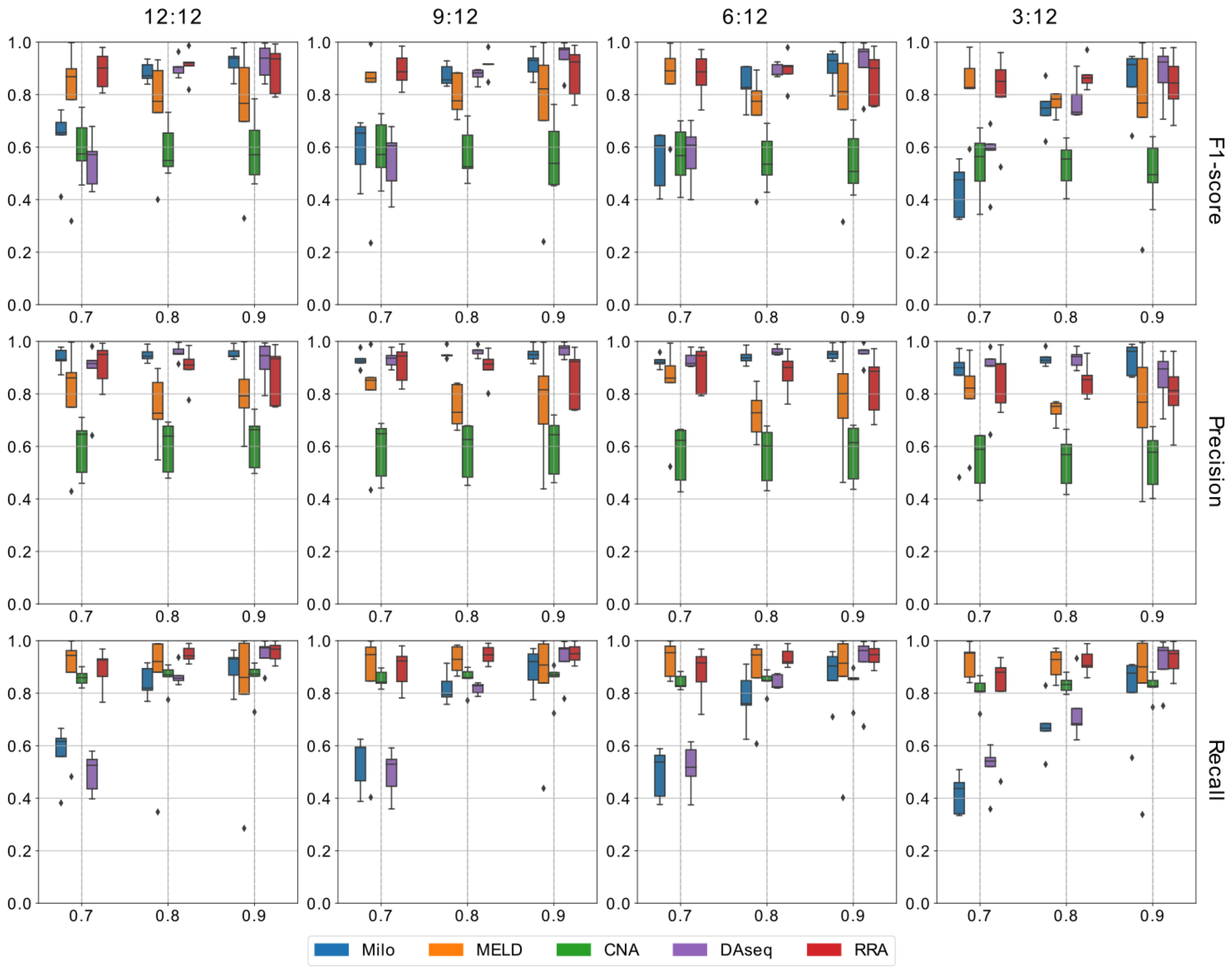


**Supplementary Figure 1: Benchmarking results of DA tools and RRA show RRA improves both accuracy and robustness compared with single DA method. Milo, MELD, CNA, DAseq, and RRA.** A probability p of 0.7, 0.8, and 0.9 is used for each synthetic dataset group within a specific subplot. Donor number $n_1$:$n_2$ means there are $n_1$ donors of condition 1, and $n_2$ donors of condition 2. Metric F1-score, precision, or recall are used for each of the three rows.

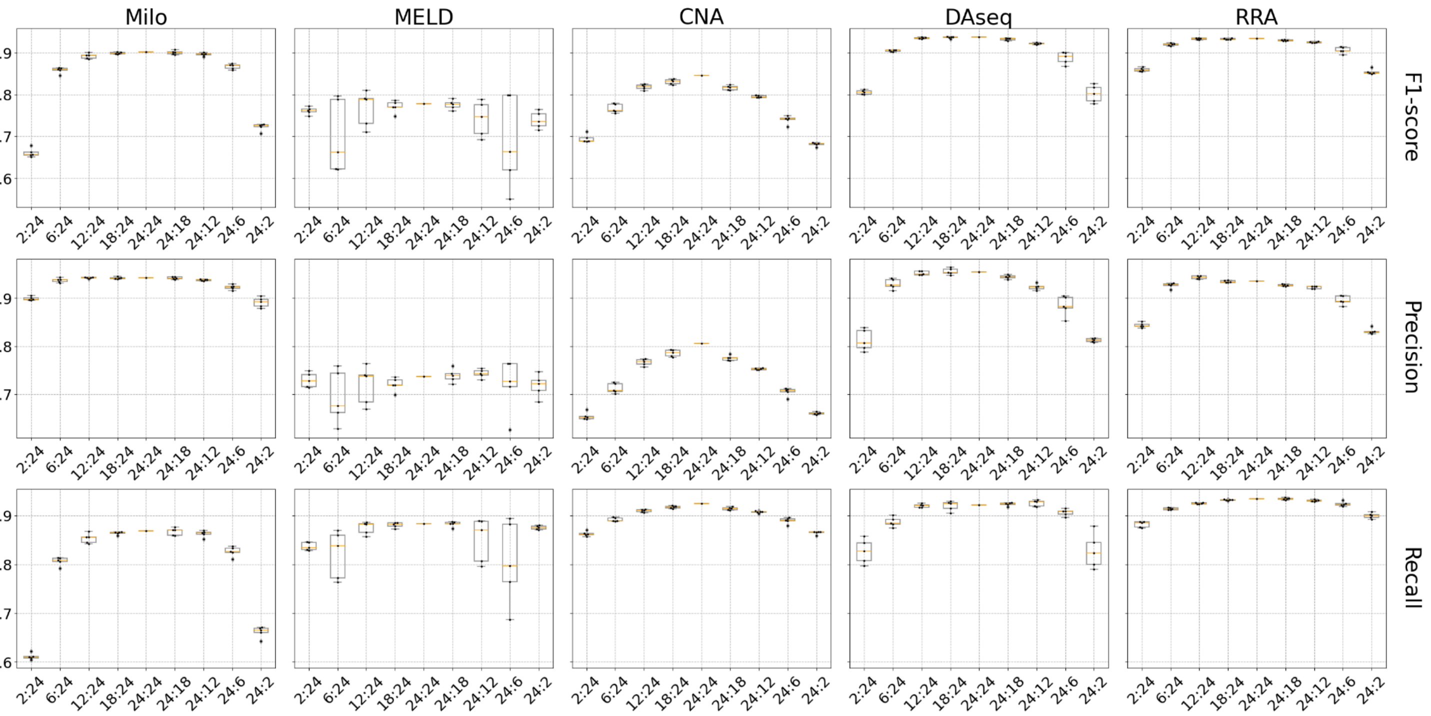


**Supplementary Figure 2: Benchmarking results of synthetic datasets with imbalanced donor numbers across conditions.** A probability p of 0.9 is used for all datasets. Donor number $n_1$:$n_2$ means there are $n_1$ donors of condition 1, and $n_2$ donors of condition 2 ($n_1$:$n_2$ = 2:24, 6:24, 12:24, 18:24, 24:24, 24:18, 24:12, 24:6, 24:2). RRA is more robust and has smaller variances compared with other single DA methods.

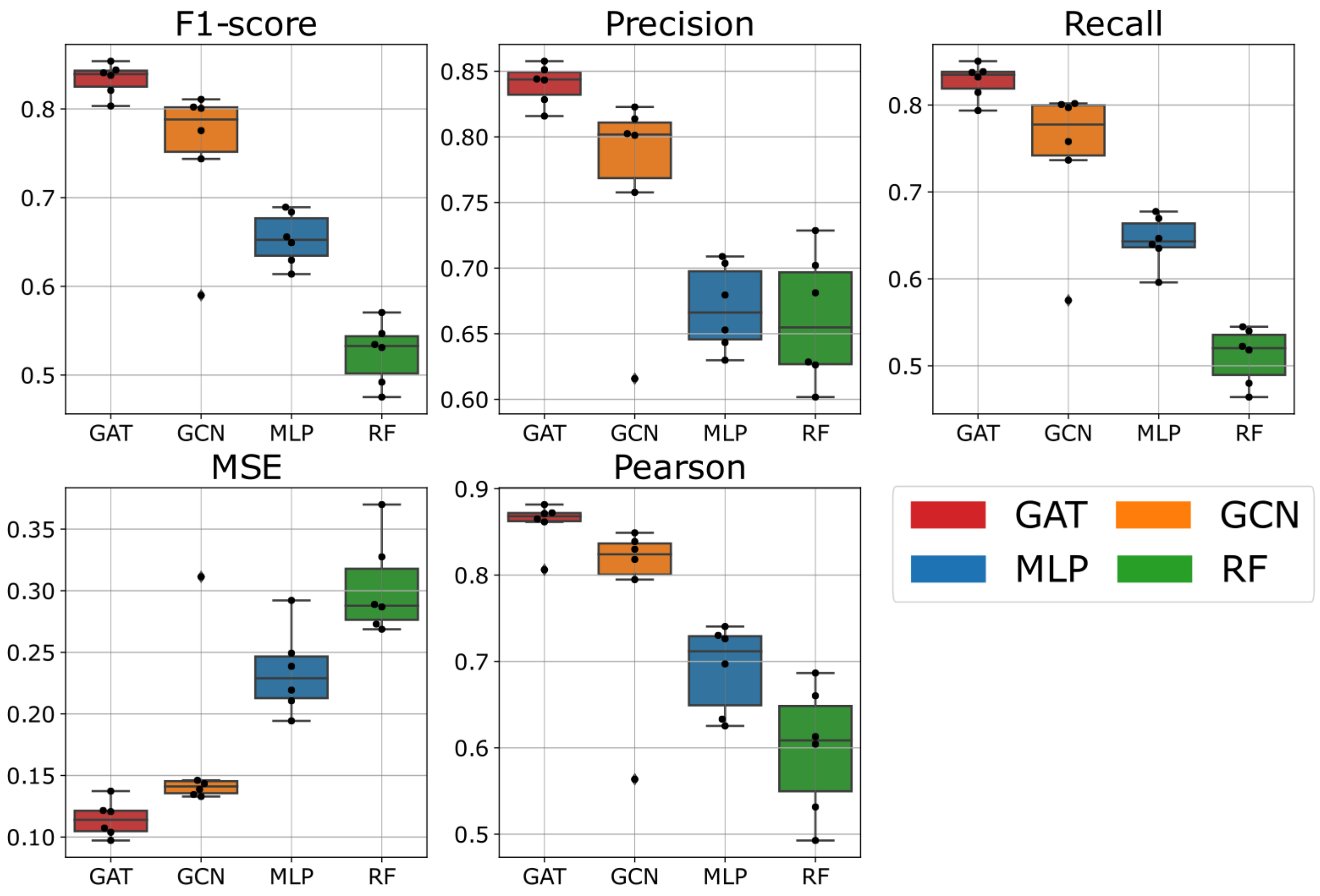


**Supplementary Figure 3: Benchmarking results of machine learning models for aggregated phenotype labels predictions using real data from** [20]**.** GAT models have superior performance over several other common machine learning models. GAT, graph attention network; GCN, graph convolutional network; MLP, multi-layer perceptron; RF, random forest; MSE, mean squared error; Pearson, Pearson correlation score.

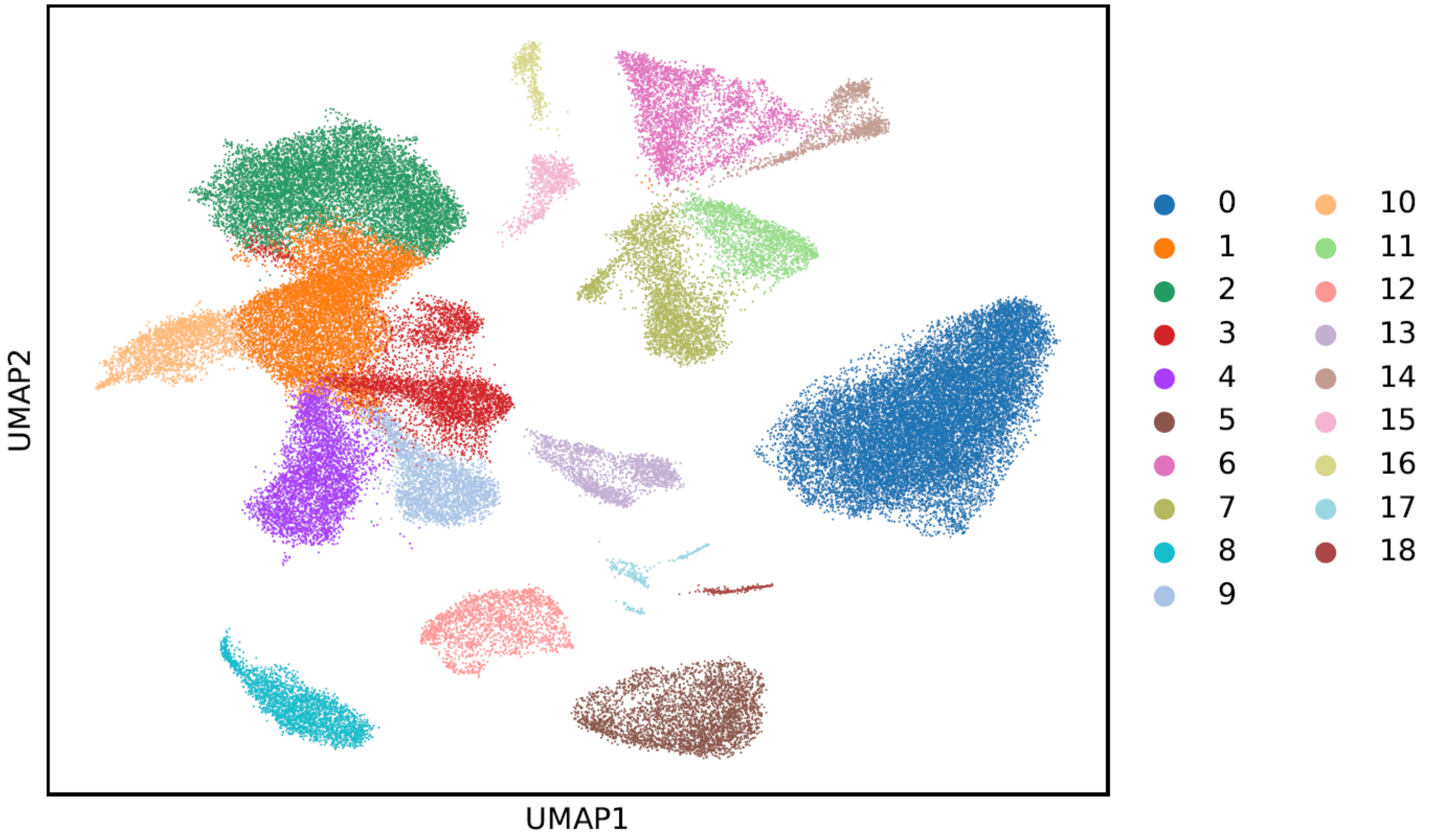


**Supplementary Figure 4: ROSMAP single-cell atlas[20] Leiden clustering.** Leiden clustering on ROSMAP single-cell RNA-seq dataset for selecting simulated ground truth for benchmark experiments. See details in **Supplementary Note 1.2.1**

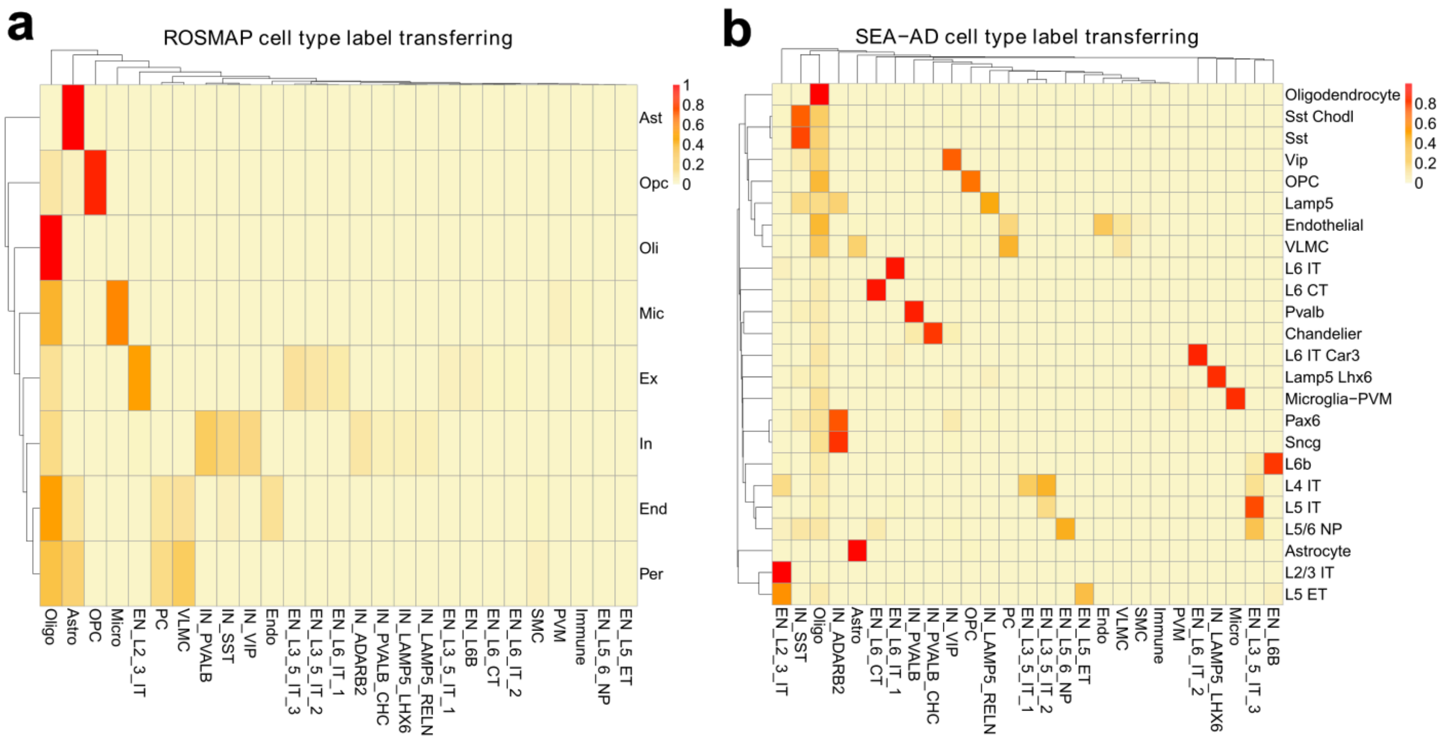


**Supplementary Figure 5: SEA-AD and ROSMAP cell type label transferring results using PsychAD cell subclasses. a,** SEA-AD original cell type labels (rows) and transferred labels (columns), and the overlapped cell number proportions. **b,** ROSMAP original cell type labels (rows) and transferred labels (columns), and proportions of the overlapped cells.

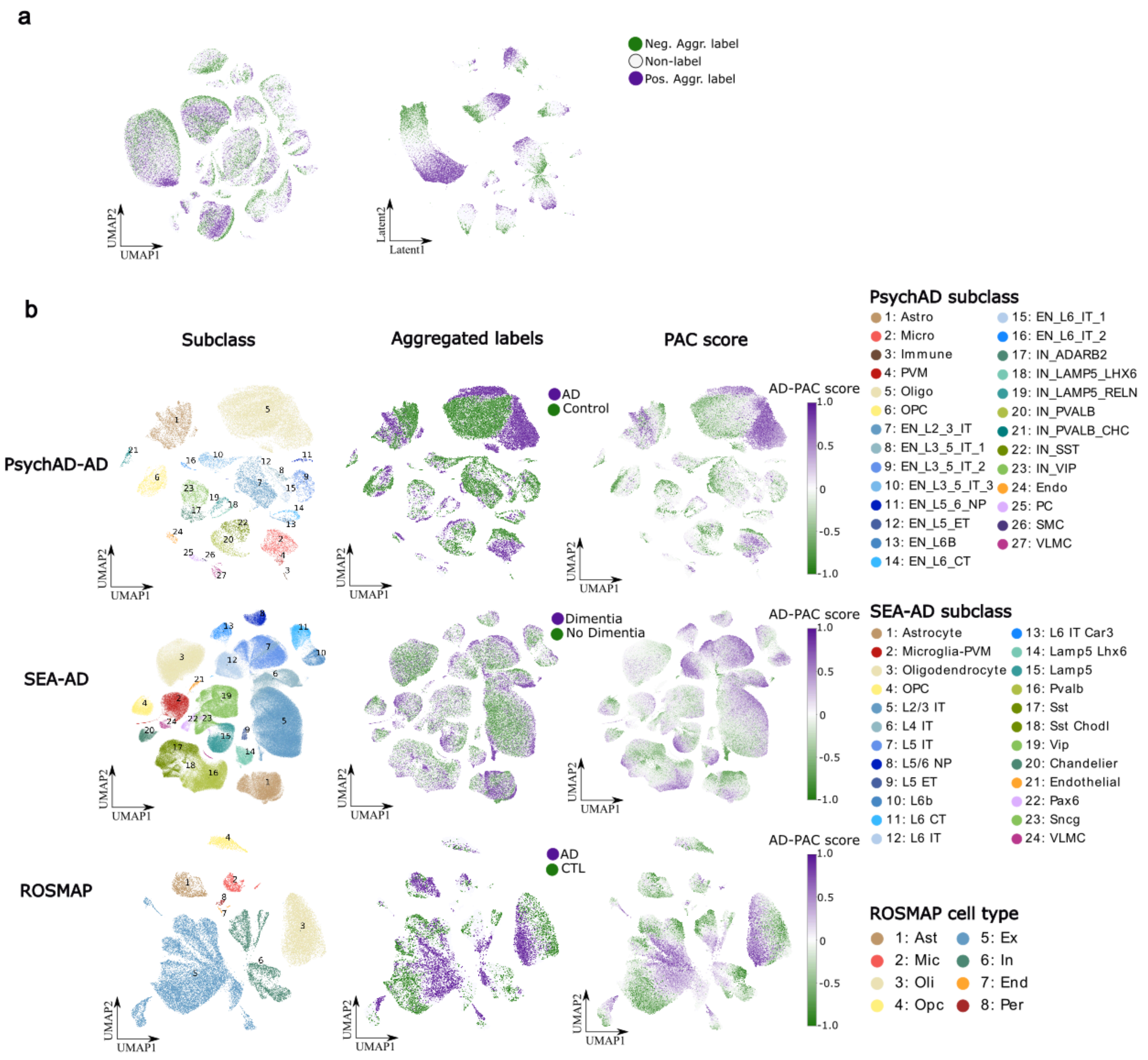


**Supplementary Figure 6. GAT separates aggregated labels in the latent space and predicts PAC scores for validation datasets. a**, UMAP of PsychAD training dataset (100 AD donors vs. 100 Control donors) (left), and the UMAP of the GAT latent space of the PsychAD training dataset (right), both colored by aggregated phenotype labels. **b**, UMAP of the three validation datasets (PsychAD held out 11 AD vs 11 Control donors, SEA-AD, ROSMAP) colored by cell types, aggregated phenotype labels, and PAC score. The aggregate phenotype label was calculated using the DA methods and the subsequent RRA algorithm as described in **Supplementary Note 1.1**.

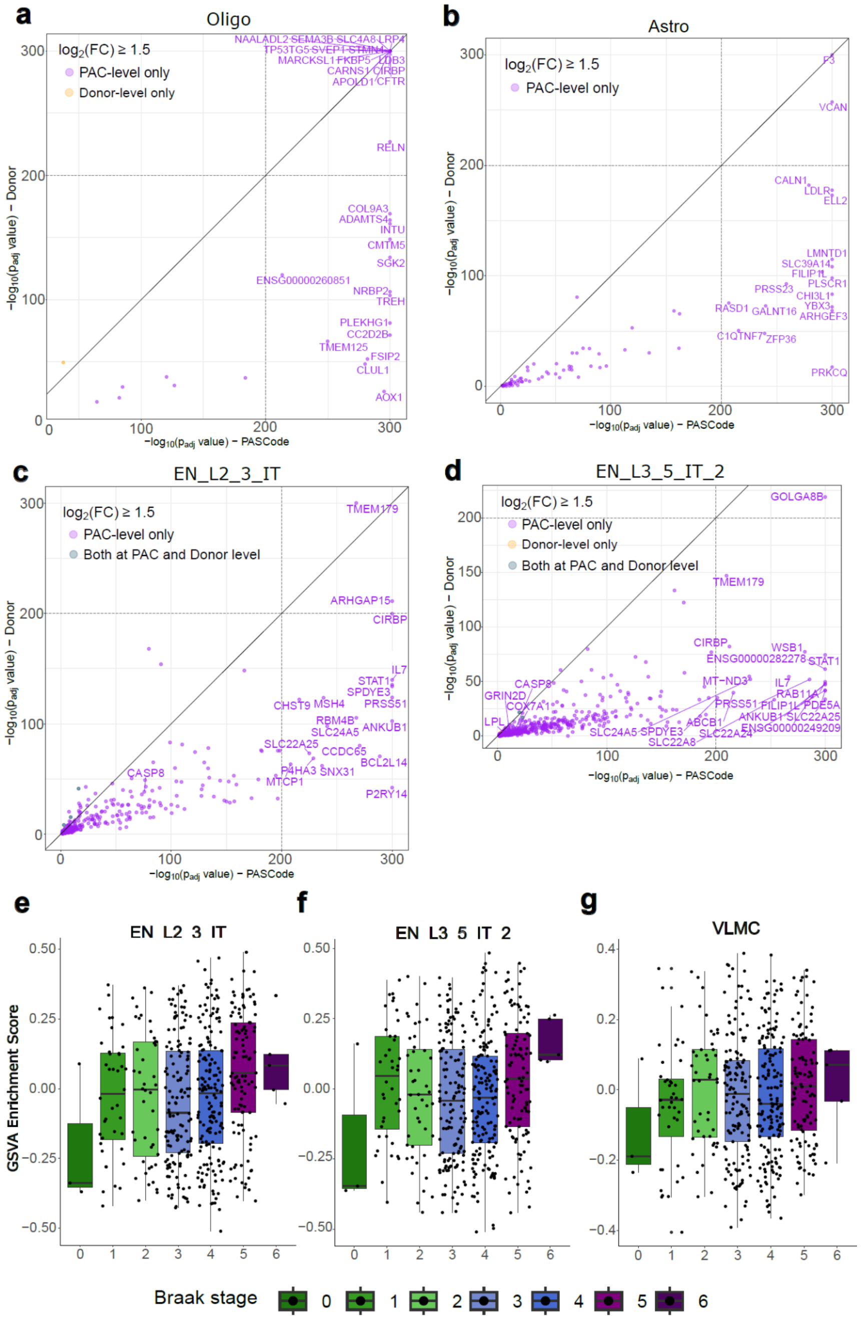

**Supplementary Figure 7: DE genes comparison between PAC-level and donor-level and GSVA enrichment of ROSMAP bulk RNAseq dataset. a-d,** Statistical significance (p-values) comparison on PAC-level versus donor-level for Oligo **(a)**, Astro **(b)**, EN_L2_3_IT **(c)**, and EN_L3_5_IT_2 **(d)**. Only genes with $log_2$ fold change bigger than 1.5 were included for comparison. Genes highlighted in orange are those that can only be found when using all cells, genes highlighted in purple are those only found when using PACs. EN_L2_3_IT **(i)**, EN_L3_5_IT_2 **(j)**, VLMC **(k)** Oligo: Oligodendrocyte; Astro: Astrocyte. **e-g**, GSVA enrichment of ROSMAP bulk RNAseq dataset[24] on upregulated DE genes in $AD\text{-}PAC^{+}$ of EN_L2_3_IT, EN_L3_5_IT_2 and VLMC significantly correlates with Braak stages of individuals within ROSMAP EN_L2_3_IT **(e)**, EN_L3_5_IT_2 **(f)**, VLMC **(g)** (Jonckheere-Terpstra trend test p-values ≤ 6.33e-4, 3.28e-2, and 7.36e-2 for EN_L2_3_IT, EN_L3_5_IT_2 and VLMC, respectively). The FPKM gene expression values were quantile normalized and batch effects removed as described in our recent work[27].

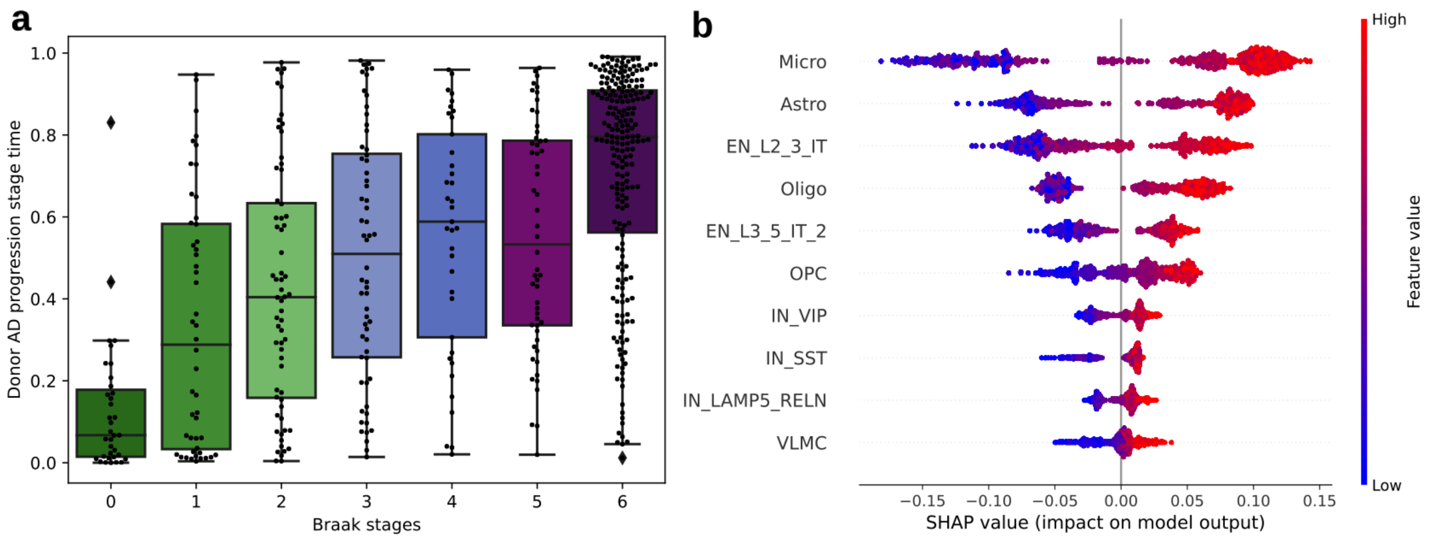


**Supplementary Figure 8: Donor AD progression stage time without AD-resilient donors, and AD progression cell type prioritization.** **a**, Donor AD progression stage time compared with Fig. 3b after removing AD-resilient donors (Jonckheere-Terpstra test p-value < $10^{-4}$.). **b**, AD progression cell type prioritization.

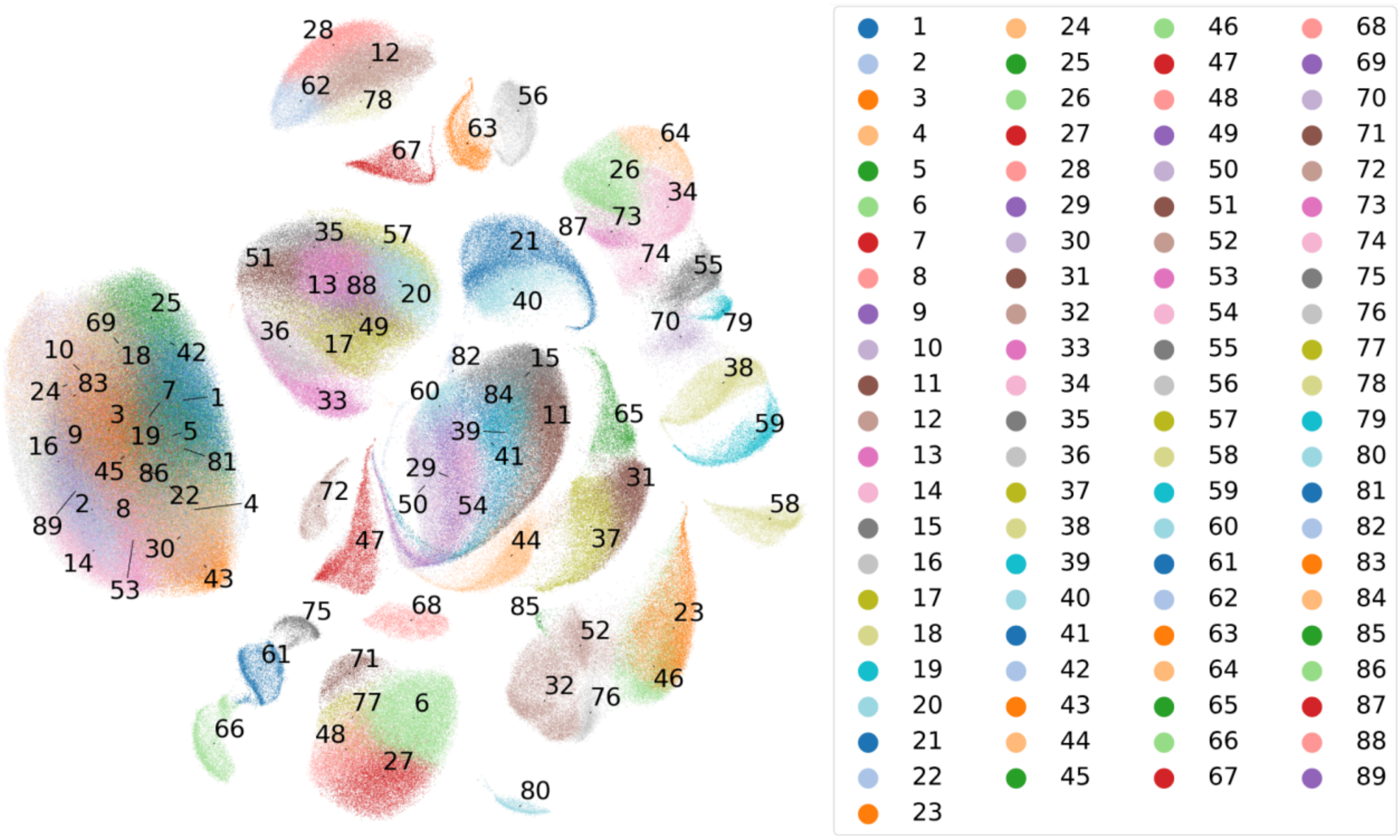

**Supplementary Figure 9: UMAP of leiden clustering of AD-Depression.** UMAP of **Fig. 2** training subset (100 AD vs. 100 Control) and **Fig. 4** training subset (63 Depression Mood vs. 63 Control) overlapping cells, colored by 89 leiden clusters.

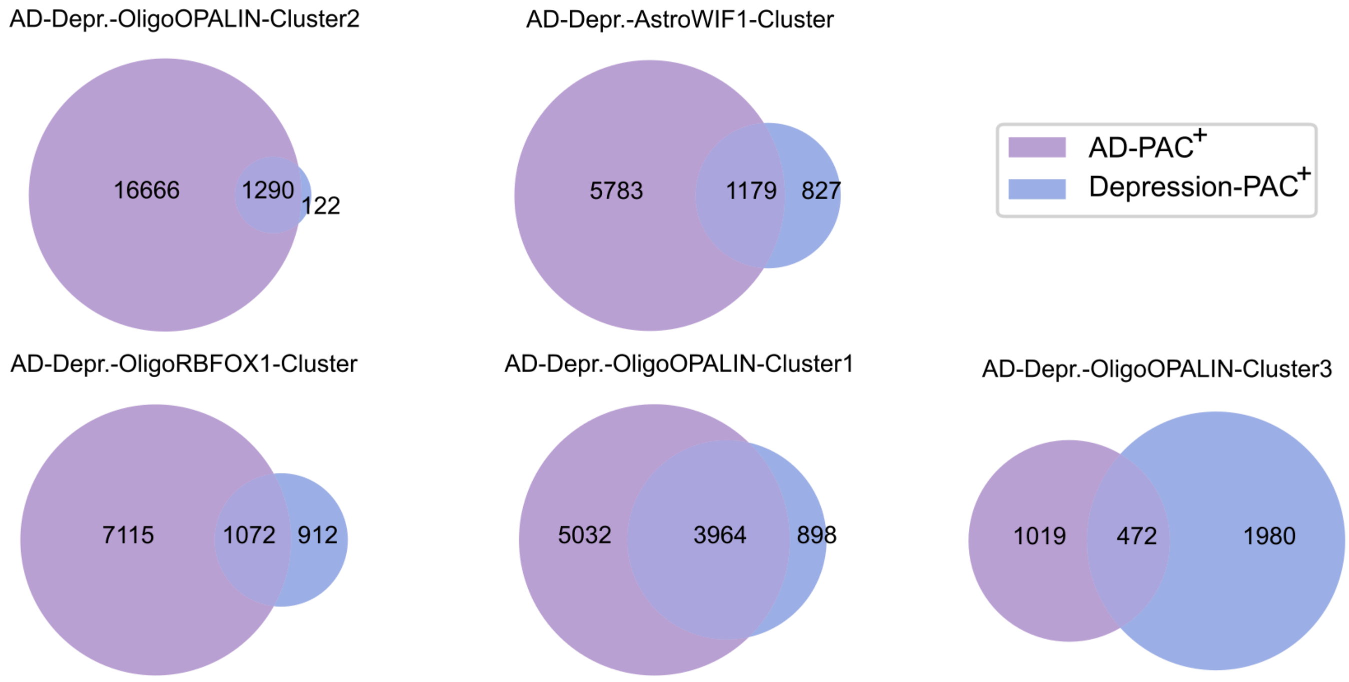


**Supplementary Figure 10: Venn diagram showing the overlapping PAC$^{+}$ numbers between AD and Depression in the significant leiden subclusters.**

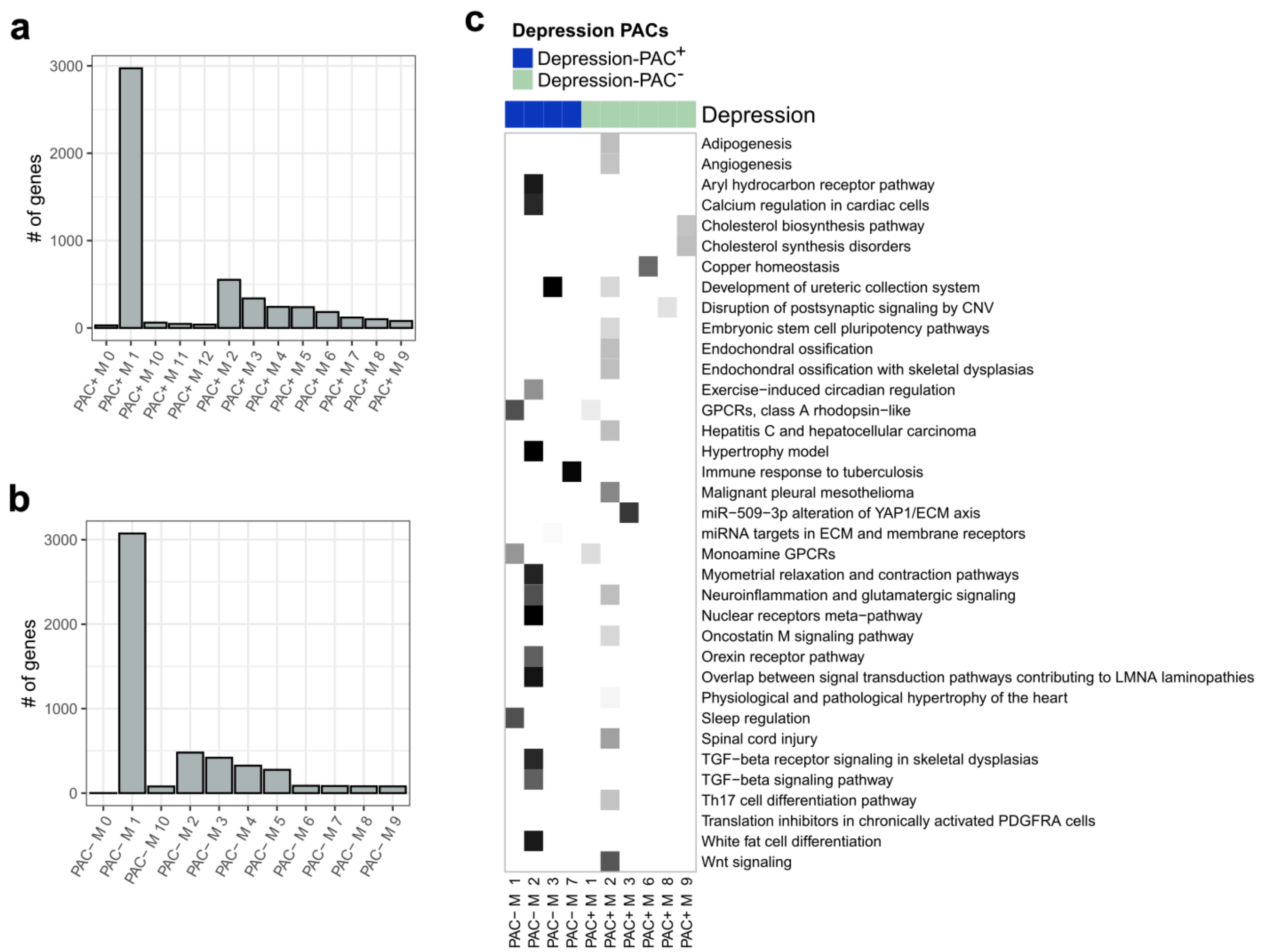


**Supplementary Figure 11. Coregulated modules. a-b,** The size (number of genes; y axis) of each module detected in $Depression\text{-}PAC^{+}$ and $Depression\text{-}PAC^{-}$ gene networks. **c,** Pathways enriched within the identified modules. The p-value of enrichment was calculated using a hypergeometric test and enrichments with an FDR < 0.1 are shown. The right annotation is color coded red and green for $Depression\text{-}PAC^{-}$ and $Depression\text{-}PAC^{+}$, respectively.

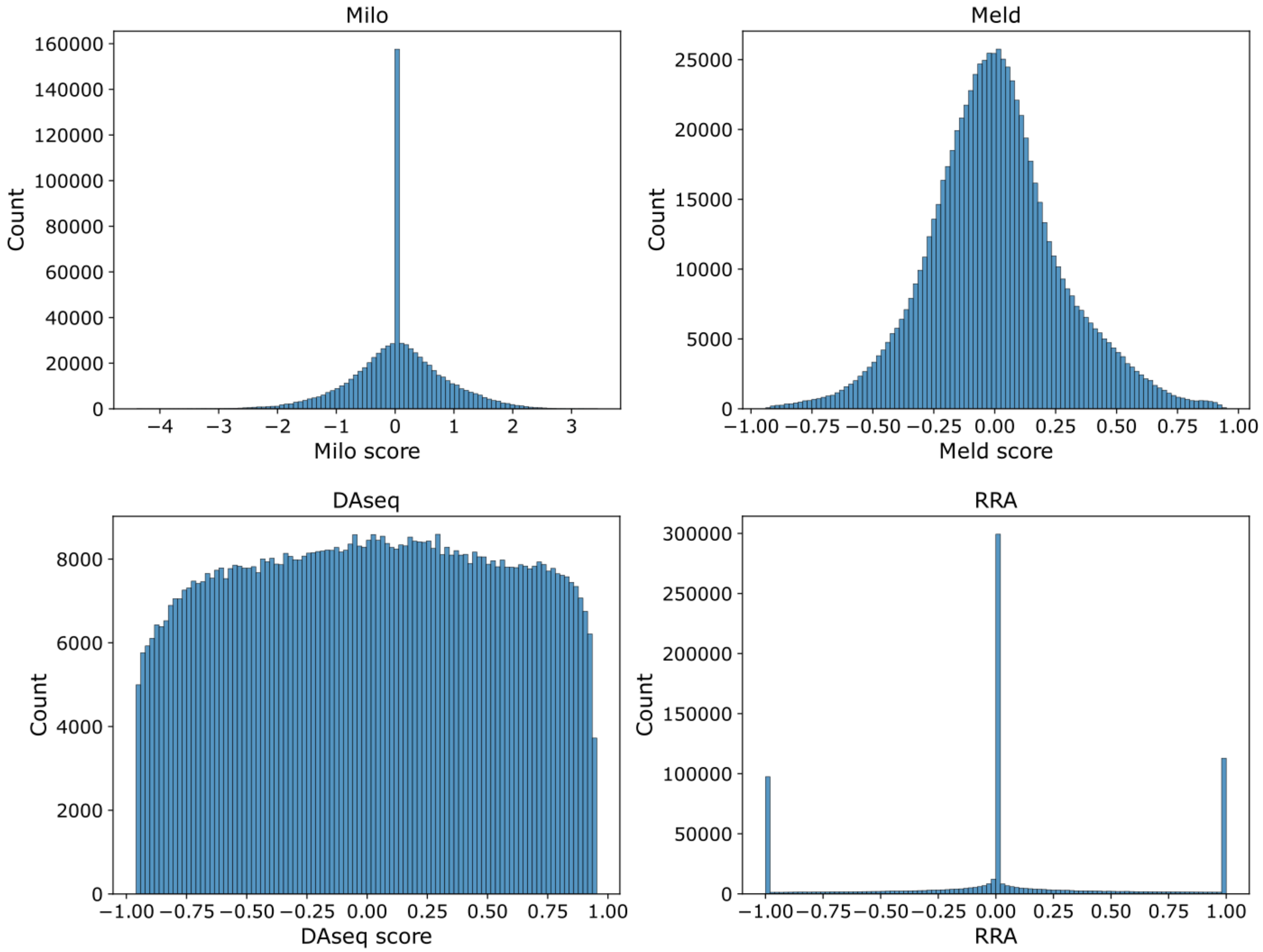


**Supplementary Figure 12.** DA methods score distribution for Milo, MELD, DAseq and RRA on **Fig. 2** training dataset (100 AD vs. 100 Control donors).

### Supplementary Tables

**Supplementary Table 1: Summary of curated differential abundance cells identification tools**

| Tool/Method | Category | Description | Whether selected for PASCode |
|---|---|---|---|
| Cydar[3] | Clustering-based | This method allocates cells to hyperspheres in the transcriptomic space, then it performs differential tests for differential abundance of each hypersphere between phenotypes. Cells within the differential abundant hyperspheres are phenotype related cells. | No |
| diffcyt[4] | Clustering-based | This method uses high-resolution clustering to group cells into a large number of small clusters. Each cluster represents cell populations or subsets, which can then be analyzed by differential testing to determine differential abundance. | No |
| Milo[7] | Clustering-free | Milo begins by constructing a cell-cell similarity graph using KNN. The neighborhood choice is refined through a sampling algorithm applied over the graph. Subsequently, statistical tests within a negative binomial (NB) generalized linear model (GLM) framework are used to identify differentially abundant neighborhoods, resulting in a spatial false discovery rate (SFDR) and log fold change (LFC) value for each neighborhood. To translate neighborhood-level results to single-cell level, for each cell, the SFDR and LFC of all the neighborhoods containing that cell are averaged and the result is assigned to that cell. The LFC values are used as the continuous measure for RRA integration. Milo's Python implementation is employed in our framework, which can be accessed at https://github.com/emdann/milopy. | Yes |
| MELD[8] | Clustering-free | MELD starts by constructing a cell-cell similarity graph using an alpha-kernel similarity function defined on the basis of k-nearest neighbors. By utilizing kernel | Yes |

| | | | |
|---|---|---|---|
| | | density estimation (KDE) methods, it defines and computes a sample-associated density estimate, quantifying the density of each sample over the cell transcriptomic state manifold. Then, based on the sample-associated density estimates, the relative likelihoods of cells belonging to a particular condition are derived. MELD's official Python implementation at https://github.com/KrishnaswamyLab/MELD is employed.<br><br>In this work, for RRA integration, we scale the likelihoods from [0,1] to [-1,1] to be treated as the continuous measure. | |
| DAseq[9] | Clustering-free | DAseq computes multiple nearest-neighbor scales for each cell from a list of k values. Each cell is then attributed a score vector, where each element is the biological state score calculated from a unique k value. A differential abundance (DA) measurement for each cell is derived from the output of a logistic regression model that takes the score vector as input. This DA measurement is within range [-1, 1], and is treated as the continuous measure for RRA integration. We employed DAseq's official R implementation from https://github.com/KlugerLab/DAseq. | Yes |
| CNA[5] | Clustering-free | CNA first uses UMAP to construct a cell-cell similarity graph, and then defines the transcriptional neighborhoods anchored at each cell. Then, CNA defines the similarity of cell i to the anchoring cell j within a transcriptional neighborhood as the probability of a random walk starting at cell i ending up at cell j after a predetermined step count. Subsequently, a sample-by-cell matrix named the neighborhood abundance matrix (NAM) is constructed. The elements of NAM represent the relative abundance of neighborhoods across samples. After performing a singular value decomposition (SVD) on NAM, a global association test with multivariate | No.<br>Benchmark experiments show relatively lower accuracy than other tools. |

| | | | |
|---|---|---|---|
| | | F-test is performed to test the association between NAM and a sample-level covariate y, which in our study is a donor-level phenotype of interest. The global association test also finds the optimal number of columns k in the left singular matrix of SVD that yields the minimal p-value. With the optimal k, a local association test is performed to derive the neighborhood coefficients for each cell, which we treat as the continuous measure to be input to RRA. We used the official python implementation of CNA from https://github.com/immunogenomics/cna. | |
| Scissor[10] | Clustering-free | Scissor takes inputs from both single-cell data and bulk sample data, and computes their correlation and then uses a regression model trained on the correlation matrix to produce single cell coefficients associated with the phenotype of interest, and the coefficients are used as DA measurement. | No.<br>Requires both bulk and single-cell data as input. |
| PENCIL[19] | Clustering-free | PENCIL uses deep learning to select DA cells and associated genes simultaneously. The same gene weights were assigned for cells with various cell types. | No.<br>The same gene weights are assigned for cells with different cell types. However, AD involves perturbation of different gene functions within different cell types. |
| PACSI[28] | Clustering-free | PACSI identifies phenotype-associated cell subpopulations by integrating scRNA-seq and bulk expression data with protein-protein interaction networks. It constructs cell signatures using highly expressed genes, calculates network-based proximity to measure cell-phenotype correlations, and assesses significance through random gene assignments. | No.<br>Requires both bulk and single-cell data as input. |

**Supplementary Table 2: Parameter choices of the selected differential abundance methods for PsychAD, ROSMAP, and SEA-AD.**

| Method | Parameter | PsychAD | ROSMAP | SEA-AD | Synthetic benchmarking |
|---|---|---|---|---|---|
| Milo | proportion (proportion of cells sampled for neighborhood indices) | 0.05 | 0.1 | 0.1 | 0.1 (default) |
| | k (number of nearest neighbors) | 15 (default) | 15 (default) | 15 (default) | 15 (default) |
| MELD | beta (a user-configurable weight parameter) | 10 | 10 | 10 | 60 (default) |
| | knn (number of nearest neighbors) | 15 | 15 | 15 | 5 (default) |
| DAseq | k (number of nearest neighbors) | 50,100,150, ..., 500 (default) | 50,100,150, ..., 500 (default) | 50,100,150, ..., 500 (default) | 50,100,150, ..., 500 (default) |

**Supplementary Table 3: Statistical tests for enrichment of three AD-related GO terms across AD vs. Control at donor and PAC levels in Microglia (Fig. 2f).**

| Mann-Whitney U rank test (pairwise comparison**) | Alzheimer's disease | | Amyloid-beta binding | | Tau-protein binding | |
|---|---|---|---|---|---|---|
| | KEGG_hsa05010 | | GO_0001540 | | GO_0048156 | |
| | Donor level | PAC level | Donor level | PAC level | Donor level | PAC level |
| **p-value** | 1.52E-80 | 7.13E-56 | 3.91E-43 | 8.42E-61 | 3.21E-79 | 6.13E-79 |
| **Difference in Mean** | 0.00377 | 0.00617 | 0.004744 | 0.01127 | 0.0106 | 0.0201 |

**AUCell estimates of each AD related enrichment terms across AD vs. Control were compared at donor and PAC level.

**Supplementary Table 4: Statistical tests to investigate the differences in enrichment across AD vs. Control, observed at the PAC level, are significantly higher than those at the Donor level in Microglia (Fig. 2f).**

| Based on Bootstrap resampling together with Mann-Whitney U rank test | Alzheimer's disease | Amyloid-beta binding | Tau-protein binding |
|---|---|---|---|
| | KEGG_hsa05010 | GO_0001540 | GO_0048156 |
| **p-value** | 0 | 0 | 0 |
| **Difference in mean at Donor level** | 0.00377 | 0.00474 | 0.01065 |
| **Difference in mean at the PAC level** | 0.00616 | 0.01127 | 0.02017 |

**Supplementary Table 5: Statistical tests for enrichment of PAC based upregulated differentially expressed genes of three cell types across AD vs. Control at donor and PAC levels within 6 cell subclasses (Extended Fig. 2c-e).**

| Mann-Whitney U rank test (pairwise comparison) | | psychAD-MSSM | | SEA-AD | | ROSMAP | |
|---|---|---|---|---|---|---|---|
| | | p-value | Difference in Mean | p-value | Difference in Mean | p-value | Difference in Mean |
| Astrocytes | Donor | 0 | 0.02479 | 0 | 0.01966 | - | - |
| | PAC | 0 | 0.07210 | 0 | 0.07998 | - | - |
| Microglia | Donor | 0 | 0.04653 | 0 | 0.03397 | 4.2e-4 | 0.00920 |
| | PAC | 0 | 0.11460 | 1.4e-198 | 0.14293 | 3.6e-5 | 0.03377 |
| Oligodendrocytes | Donor | 0 | 0.02383 | 0 | 0.02655 | 2.23e-23 | 0.00763 |
| | PAC | 0 | 0.07810 | 0 | 0.09901 | 4.1e-133 | 0.04022 |
| EN_L2_3_IT | Donor | 0 | 0.00683 | 0 | 0.0109 | 1.6e-49 | 0.0071 |
| | PAC | 0 | 0.02432 | 0 | 0.03122 | 1.6e-161 | 0.02756 |
| EN_L3_5_IT_2 | Donor | 4.8e-237 | 0.01259 | 0 | 0.01928 | - | - |
| | PAC | 0 | 0.04620 | 4.6e-160 | 0.04126 | - | - |
| VLMC | Donor | 7.2e-34 | 0.02263 | - | - | - | - |
| | PAC | 5.5e-146 | 0.13925 | - | - | - | - |

**Supplementary Table 6: Statistical tests to inquire the differences in upregulated differentially expressed gene enrichment across AD vs. Control observed at PAC level is significantly higher than that at the Donor level (Extended Fig 2c-e) within 6 cell subclasses.**

| Based on Bootstrap resampling together with Mann-Whitney U rank test | | Astro | Micro | Oligo | L2-3IT | L3-5IT_2 | VLMC |
|---|---|---|---|---|---|---|---|
| **psychAD-MSSM** | **p-value** | 0 | 0 | 0 | 0 | 0 | 0 |
| | **Difference in Mean at donor level** | 0.02479 | 0.04655 | 0.02383 | 0.00683 | 0.01259 | 0.02261 |
| | **Difference in Mean at PAC level** | 0.07209 | 0.11462 | 0.07810 | 0.02432 | 0.04618 | 0.13923 |
| **SEA-AD** | **p-value** | 0 | 0 | 0 | 0 | 0 | - |
| | **Difference in Mean at donor level** | 0.01967 | 0.0339 | 0.02655 | 0.0109 | 0.01928 | - |
| | **Difference in Mean at PAC level** | 0.07997 | 0.1429 | 0.0990 | 0.03122 | 0.04125 | - |

| | | | | | | | |
|---|---|---|---|---|---|---|---|
| **ROSMAP** | **p-value** | - | - | 0 | 0 | - | - |
| | **Difference in Mean at donor level** | - | - | 0.00761 | 0.0071 | - | - |
| | **Difference in Mean at PAC level** | - | - | 0.04023 | 0.0276 | - | - |

### Supplementary Data

**Supplementary Data 1.** Differential gene analysis for cell type AD related genes in Fig. 2 and Extended Fig. 2

**Supplementary Data 2.** Genes and trajectory analysis to compare AD-resilient and AD-strict donors in Fig. 3 and Extended Fig. 3

**Supplementary Data 3.** Depression and AD associated PACs analysis in Fig. 4 and Extended Fig. 4

**Supplementary Data 4.** Gene module analysis on Astrocytes PACs in Fig. 5

**Supplementary Data 5.** PAC numbers for AD, WeightGain/Sleep/Suicide, Weightloss/PMA, and Depression/Mood in Fig. 6

**Supplementary Data 6.** Average PAC scores and SHAP values across donors in each subclass for AD, WeightGain/Sleep/Suicide, Weightloss/PMA, and Depression/Mood in Fig. 6

**Supplementary Data 7.** PAC-based DEGs for AD, WeightGain/Sleep/Suicide, Weightloss/PMA, and Depression/Mood in Fig. 6
